# Multi-ancestry genetic study in 5,876 patients identifies an association between excitotoxic genes and early outcomes after acute ischemic stroke

**DOI:** 10.1101/2020.10.29.20222257

**Authors:** Laura Ibanez, Laura Heitsch, Caty Carrera, Fabiana H.G. Farias, Rajat Dhar, John Budde, Kristy Bergmann, Joseph Bradley, Oscar Harari, Chia-Ling Phuah, Robin Lemmens, Alessandro A. Viana Oliveira Souza, Francisco Moniche, Antonio Cabezas-Juan, Juan Francisco Arenillas, Jerzy Krupinksi, Natalia Cullell, Nuria Torres-Aguila, Elena Muiño, Jara Cárcel-Márquez, Joan Marti-Fabregas, Raquel Delgado-Mederos, Rebeca Marin-Bueno, Alejandro Hornick, Cristofol Vives-Bauza, Rosa Diaz Navarro, Silvia Tur, Carmen Jimenez, Victor Obach, Tomas Segura, Gemma Serrano-Heras, Jong-Won Chung, Jaume Roquer, Carol Soriano-Tarraga, Eva Giralt-Steinhauer, Marina Mola-Caminal, Joanna Pera, Katarzyna Lapicka-Bodzioch, Justyna Derbisz, Antoni Davalos, Elena Lopez-Cancio, Lucia Muñoz, Turgut Tatlisumak, Carlos Molina, Marc Ribo, Alejandro Bustamante, Tomas Sobrino, Jose Castillo-Sanchez, Francisco Campos, Emilio Rodriguez-Castro, Susana Arias-Rivas, Manuel Rodríguez-Yáñez, Christina Herbosa, Andria L. Ford, Antonio Arauz, Iscia Lopes-Cendes, Theodore Lowenkopf, Miguel A. Barboza, Hajar Amini, Boryana Stamova, Bradley P. Ander, Frank R Sharp, Gyeong Moon Kim, Oh Young Bang, Jordi Jimenez-Conde, Agnieszka Slowik, Daniel Stribian, Ellen A. Tsai, Linda C. Burkly, Joan Montaner, Israel Fernandez-Cadenas, Jin-Moo Lee, Carlos Cruchaga

**Affiliations:** Department of Psychiatry, Washington University School of Medicine, 660 S. Euclid Avenue, Saint Louis (63110), Missouri, US; NeuroGenomics and Informatics, Washington University School of Medicine, 425 S. Euclid Avenue, Saint Louis (63110), Missouri, US; Department of Neurology, Washington University School of Medicine, 660 S. Euclid Avenue; Campus Box 8111; Saint Louis (63110), Missouri, US; Emergency Medicine, Washington University School of Medicine, 660 S. Euclid Avenue; Campus Box 8072; Saint Louis (63110), Missouri, US; Stroke Unit, Vall d’Hebron University Hospital, Universitat de Barcelona, Passeig de la Vall d’Hebron, 1198; Barcelona (08035), Spain; Hope Center for Neurological Disorders, Washington University School of Medicine, 660 S. Euclid Avenue; Campus Box 8111; Saint Louis (63110), Missouri, US; The Charles F. and Joanne Knight Alzheimer Disease Research Center, Washington University School of Medicine, 4488 Forest Park Avenue; Saint Louis (63110), Missouri, US; Department of Neuroscience, Katholieke Universiteit Leuven, Campus Gasthuisberg O&N2; Herestraat 49 box 1021; Leuven (BE-3000), Belgium; Department of Neurology, School of Medical Sciences, University of Campinas (UNICAMP), R. Tessalia Viera de Camargo, 126; Cidade Universitaria, Campinas (13083-887), Brazil; Brazilian Institute of Neuroscience and Neurotecnology (BRAINN), R. Tessalia Viera de Camargo, 126; Cidade Universitaria, Campinas (13083-887), Brazil; Department of neurology, Hospital Virgen del Rocio, University of Seville, Avenida Manuel Siurot, s/n; Seville (41013), Spain; Hospital Virgen de la Macarena, University of Seville, Calle Dr. Fedriani, 3; Seville (41009), Spain; Department of Neurology, Hospital Clinico Universitario Valladolid, Valladolid University, Avenida Ramon y Cajal, 3; Valladolid (47003), Spain; Department of Neurology, Mutua Terrassa University Hospital, Universitat de Barcelona, Plaça del Dr. Robert, 5; Terrassa (08221), Spain; Fundacio Docencia i Recerca Mutua Terrassa, Universitat de Barcelona, Carrer Sant Antoni, 19; Terrassa (08221), Spain; Department of Neurology, Hospital de la Santa Creu i Sant Pau, Universitat Autonoma de Barcelona, Carrer de Sant Quinti, 89; Barcelona (08041), Spain; Department of Neurology, Southern Illinois Healthcare Memorial Hospital of Carbondale, 405 W Jackson Street, Carbondale (62901), Illinois, US; Department of Biology, Universitat de les Illes Balears, Carretera de Valldemossa, km 7,5, Palma (07122), Spain; Department of Neurology, Hospital Universitari Son Espases, Universitat de les Illes Balears, Carretera de Valldemossa, 79, Palma (07120), Spain; Department of Neurology, Hospital Clinic de Barcelona, Universitat de Barcelona, Carrer Villarroel, 170, Barcelona (08036), Spain; Research Unit, Complejo Hospitalario Universitario de Albacete. Calle Laurel s/n. Albacete (02008), Spain; Department of Neurology, Samsung Medical Center, 81 Irwon-ro, Gangnam-gu, Seoul, South Korea; Neurovascular Research Group, Institut Hospital del Mar de Investigacions Mediques, Passeig Maritim, 25-29, Barcelona (08003), Spain; Department of Surgical Sciences, Orthopedics, Uppsala University, Uppsala, 75185, Sweden; Department of Neurology, Jagiellonian University, Golebia, 24, Krakow(31-007), Poland; Department of Neurology, Hospital Germans Trias i Pujol, Universitat Autonoma de Barcelona, Carretera de Canyet, s/n; Badalona (08916), Spain; Department of Neurology, Hospital Universitario Central de Asturias, Oviedo, Spain; Department of Neurology, Sahlgrenska University Hospital, University of Gothenburg, Bla straket, 5; Gothenburg (413 45), Sweden; Department of Clinical Neuroscience, Institute of Neuroscience and Physiology, Sahlgrenska Academy at University of Gothenburg, Gothenburg, Sweden; Clinical Neurosciences Research Laboratory, Health Research Institute of Santiago de Compostela (IDIS), Avda. Travesa da Choupana s/n; Santiago de Compostela (15706), Spain; Department of Radiology, Washington University School of Medicine, 660 S. Euclid Avenue, Saint Louis (63110), Missouri, US; Instituto Nacional de Neurologia y Neurocirurgia de Mexico, Avenida Insurgentes Sur 3877, Ciudad de Mexico (14269), Mexico; Department of Neurology, Providence St. Vincent Medical Center, 9205 SW Barnes Rd, Portland (97225), Oregon, US; Neurosciences Department, Hospital Rafael A. Calderon Guardia, Avenidas 7 y 9, calles 15 y 17, Aranjuez, San José, Costa Rica; Department of Neurology and MIND Institute, University of California at Davis, 2825 5th street, Sacramento (95817), California, US; Department of Neurology, Helsinki University Hospital, Haartmaninkatu 4 Rakennus 1, Helsinki (00290), Finland; Translational Biology, Biogen, Inc, 115 Brodway, Cambridge (02142), Massachusetts, US; Genetics and Neurodevelomental Disease Research Unit, Biogen, Inc, 115 Brodway, Cambridge (02142), Massachusetts, US; Department of Biomedical Engineering, Washington University School of Medicine, 660 S. Euclid Avenue, Saint Louis (63110), Missouri, US; Stroke and Cerebrovascular Center, Washington University School of Medicine, One Barnes-Jewish Hospital Plaza, Saint Louis (63110), Missouri, US; Department of Genetics, Washington University School of Medicine, 4515 McKinley Ave, Saint Louis (63110), Missouri, US

**Keywords:** Ischemic Stroke, Neuroprotection, Genetics, NIHSS

## Abstract

During the first hours after stroke onset neurological deficits can be highly unstable: some patients rapidly improve, while others deteriorate. This early neurological instability has a major impact on long-term outcome. Here, we aimed to determine the genetic architecture of early neurological instability measured by the difference between NIH stroke scale (NIHSS) within six hours of stroke onset and NIHSS at 24h (ΔNIHSS). A total of 5,876 individuals from seven countries (Spain, Finland, Poland, United States, Costa Rica, Mexico and Korea) were studied using a multi-ancestry meta-analyses. We found that 8.7% of ΔNIHSS variance was explained by common genetic variations, and also that early neurological instability has a different genetic architecture than that of stroke risk. Seven loci (2p25.1, 2q31.2, 2q33.3, 4q34.3, 5q33.2, 6q26 and 7p21.1) were genome-wide significant and explained 2.1% of the variability suggesting that additional variants influence early change in neurological deficits. We used functional genomics and bioinformatic annotation to identify the genes driving the association from each loci. eQTL mapping and SMR indicate that *ADAM23* (log Bayes Factor (LBF)=6.34) was driving the association for 2q33.3. Gene based analyses suggested that *GRIA1* (LBF=5.26), which is predominantly expressed in brain, is the gene driving the association for the 5q33.2 locus. These analyses also nominated *PARK2* (LBF=5.30) and *ABCB5* (LBF=5.70) for the 6q26 and 7p21.1 loci. Human brain single nuclei RNA-seq indicates that the gene expression of *ADAM23* and *GRIA1* is enriched in neurons. *ADAM23*, a pre-synaptic protein, and *GRIA1*, a protein subunit of the AMPA receptor, are part of a synaptic protein complex that modulates neuronal excitability. These data provides the first evidence in humans that excitotoxicity may contribute to early neurological instability after acute ischemic stroke.

**RESEARCH INTO CONTEXT:** *Evidence before this study:* No previous genome-wide association studies have investigated the genetic architecture of early outcomes after ischemic stroke.

*Added Value of this study:* This is the first study that investigated genetic influences on early outcomes after ischemic stroke using a genome-wide approach, revealing seven genome-wide significant loci. A unique aspect of this genetic study is the inclusion of all of the major ethnicities by recruiting from participants throughout the world. Most genetic studies to date have been limited to populations of European ancestry.

*Implications of all available evidence:* The findings provide the first evidence that genes implicating excitotoxicity contribute to human acute ischemic stroke, and demonstrates proof of principle that GWAS of acute ischemic stroke patients can reveal mechanisms involved in ischemic brain injury.

## Introduction

Stroke is the second most common cause of death and the most common cause of disability, worldwide.^1^ Ischemic stroke, the most common subtype^2^, is caused by the occlusion of an artery in the brain, resulting in the abrupt development of cerebral ischemia and neurological deficits.^3^ During the first hours after stroke onset, neurological deficits can be highly unstable with some patients demonstrating rapid deterioration, while others rapidly improve.^4^ In fact, early change in neurological deficits have a major influence on long-term outcome. NIH stroke scale (NIHSS) changes from baseline (within 6 hours of stroke onset) to 24 hours after acute ischemic stroke (ΔNIHSS) have a significant and independent association with favorable 90-day outcome, accounting for more than 30% of the explained variance.^4-6^ A number of mechanisms are thought to account for these early changes including fibrinolysis and reperfusion, hemorrhagic transformation, etiology, and endogenous neuroprotective mechanisms.^7-14^

Prior genome wide association studies (GWAS), mostly in populations of European descent, have identified numerous loci associated with stroke risk. In 2018, the MEGASTROKE consortia performed one of the largest GWAS to date, combining most of the available GWAS for stroke risk in a unique multi-ancestry meta-analysis including 67,162 cases and 454,450 controls. This analysis led to the discovery of 22 novel loci, bringing the total stroke risk loci to 32. Many loci were previously linked to other vascular traits (blood pressure, cardiac phenotypes, venous thromboembolism); while others had no obvious connection with stroke, warranting further investigation to identify potentially novel mechanisms.^15^ A similar approach, used to decipher the genetics of long term disability after ischemic stroke in 6,165 non-Hispanic Whites, identified one locus that was been not replicated so far.^16,17^ However, to date there have been no genetic studies examining early neurological change after ischemic stroke.

To our knowledge, the Genetics of Early Neurological InStability after Ischemic Stroke (GENISIS) is the largest well-characterized study for early outcomes quantified by ΔNIHSS.^18^ To increase the power to detect genetic associations, our study recruited patients from multiple diverse ancestry groups. We leveraged the GENISIS cohort using ΔNIHSS as a quantitative phenotype, to identify novel variants, genes and pathways associated with early neurological instability after ischemic stroke.

## Materials and Methods

### Study Design

A detailed description of the acute ischemic stroke patients recruited from 21 sites from seven countries throughout the world, has been published elsewhere.^6^ Briefly, adult acute ischemic stroke patient with measurable deficit on the NIHSS that presented within 6 hours of stroke onset (or last known normal) were enrolled in the study after obtaining informed consent, including patients treated with tPA. Patients who underwent a thrombectomy, were enrolled in other treatment trials, or for whom consent and/or a blood sample could not be obtained were excluded. Demographics, co-morbidities, acute treatment variables, imaging data and TOAST classification were collected.

To accommodate the difference in the genetic architecture intrinsic to the country of origin, we performed a three-stage analysis (Figure 1A). First, we used an additive model to perform a GWAS in each country individually, except for the United States, where the population was stratified into European and African ancestry cohorts. We then performed a fixed effects meta-analyses within the same ethnic cohorts. Finally, we used a multi-ancestry Bayesian meta-analysis to collapse all the ethnic backgrounds. Unlike a fixed effect meta-analysis, the Bayesian approach is able to account for population structure differences.^19^ Genetic loci that passed multiple test correction, a threshold set at Log Bayes Factor (LBF) > 5, were annotated using bioinformatics tools to identify the gene driving the genetic signal (Figure 1B). We used functional annotation, multi-tissue expression quantitative trait loci (eQTL) data, and summary-data-based Mendelian randomization (SMR) to map the genome-wide to specific genes. Single nuclei-RNA-seq data derived from cortex samples was used to determine potential correlation between the transcripts of the identified genes and determine in which brain cell types the genes are expressed.^20^

**Figure 1.**
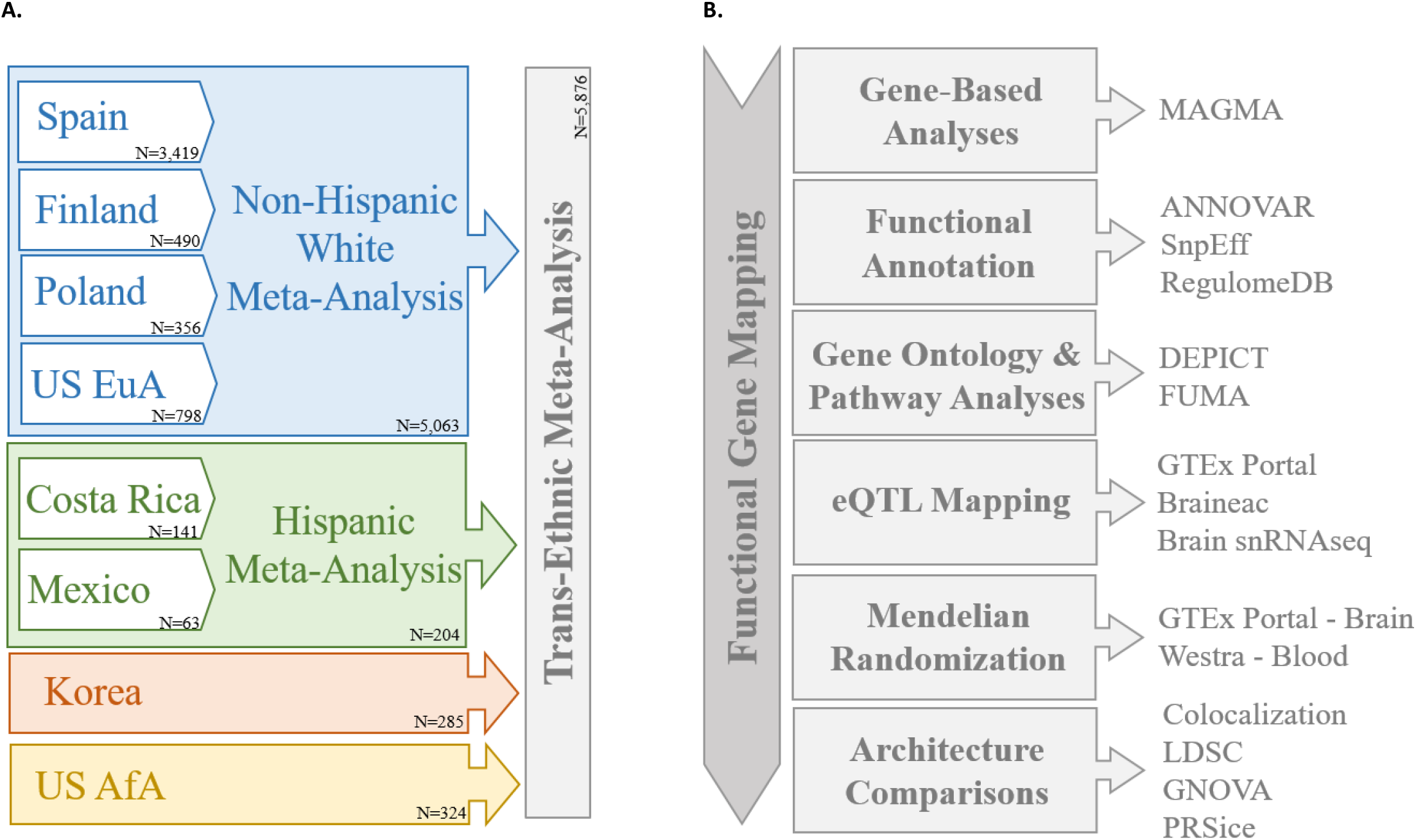
Study design. Summarized description of **A**. the multi-step approach used to account for the genetic heterogeneity intrinsic to the multi-ancestry nature of the GENISIS study. We performed single variant analysis in each of the participating countries separately. Then we meta-analyzes all the non-Hispanic whites (blue) and Hispanic (green) ethnicities. Finally, we analyzed the non-Hispanic whites, Hispanics, Korea (orange) and US participants with African descent (US AfA – yellow) using a Bayesian model. The variants with genome-wide significant or suggestive results were annotated using **B**. sequential steps to elucidate the gene driving the association. We performed gene-based and pathway analyses, we collected the information available in publicly available datasets and we performed Mendelian randomization. We also performed genetic architecture overlap tests to test if there was any overlap with known risk factors.

The study was approved by the Institutional Review Boards at every participating site. Written informed consent was obtained from all participants or their family members. All research was performed according to the approved protocols and consents.

### Genotyping

All participants were genotyped using Illumina SNP array technology. Samples were genotyped in seven batches during the GENISIS recruitment (see Supplementary Methods). Genotyping quality control and imputation were performed separately for each genotyping round using SHAPEIT^21^ and IMPUTE2.^22^ For each genotyping batch, SNPs with a call rate lower than 98% and autosomal SNPs that were not in Hardy-Weinberg equilibrium (P<1×10^−06^) were removed from the dataset. The X chromosome SNPs were used to determine sex based on heterozygosity rates, and samples with discordant inferred sex and reported sex were removed. Only samples with call rate greater than 98% were considered to pass quality control. Finally, the genotype batches were merged in a single file to perform the analyses.

Additional QC was performed in the merged dataset. We tested pairwise genome-wide estimates of proportion identity-by-descent, the presence of unexpected duplicates, and cryptically relatedness (PI-HAT>0.30). Of the pairs of these samples flagged, the sample with higher genotyping rate was kept for downstream analysis. Principal component analysis (PCA) was performed using HapMap as an anchor to remove ethnic outliers and keep the populations as homogeneous as possible for each of the participant countries. Principal components were also used to cluster and identify ancestry populations for US participants with European descent (EuA) and African-American descent (AfA). Samples outside two standard deviations from the center of the Non-Hispanic White or the Asian cluster were considered outliers for Spain, Finland and Poland. We confirmed the ethnicity of the AfA and Hispanic populations, however, due to the genetic heterogeneity present in these populations we did not remove the samples outside two standard deviations from the mean.

### Analysis of variance

We used genome-wide complex traits analysis (GCTA) to determine the heritability of ΔNIHSS.^23^ GCTA estimates the amount of phenotypic variance in a given complex trait explained by all the SNPs and fits the effects of these SNPs as random effects in a linear mixed model. Because it relies on a large, homogeneous populations for accurate results, we only included the individuals with non-Hispanic White ancestry.

### Single Variant Analyses

To mitigate the effects of genetic heterogeneity due to the diverse ancestry of participants enrolled in the GENISIS study, we used a multi-step study design (Figure 1A). First, we performed single variant analyses each participant country separately. We tested the association of SNPs across the genome with ΔNIHSS using an additive linear model with PLINK 1.9.^24^ Sex, age, and the two Principal Components calculated for each population were included in the model. Additional covariates include the SNP genotyping batch, TOAST classification (using dummy variables to incorporate all subtypes), and baseline NIHSS to adjust for stroke severity. Although baseline NIHSS was used to calculate ΔNIHSS, it does not fully explain the observed variance in ΔNIHSS; further, there is no multicollinearity between these two variables, permitting their inclusion in the model.^25^ Fifty four percent of study participants were treated with tPA, however treatment was not included as covariate. While tPA influences ΔNIHSS^6^, it has little impact on the genetic architecture of ΔNIHSS. The inclusion of tPA as covariate lead to highly correlated results when compared to a model without tPA (r^2^=0.96, Supplementary Figure 1). Second, we meta-analyzed the populations with similar ethnic backgrounds using with fixed effect meta-analyses using METAL.^26^ We performed two meta-analyses, one for the non-Hispanic Whites (Spain, Finland, Poland, and United States EuA) and one for the Hispanics (Costa Rica and Mexico). Finally, we analyzed the four available ethnicities non-Hispanic Whites (meta-analysis), Hispanics (meta-analysis), Asians (Korea) and African Americans (United States AfA) using MANTRA, a Bayesian-based multi-ancestry meta-analyses.^19^ Log Bayes Factor (LBF) greater than 5 was considered to be genome wide significant after multiple test correction.

### Functional Annotation

We annotated all the variants with suggestive associations (LBF>4) with ANNOVAR^27^ and SnpEff^28^ to identify the nearest gene and to determine if any variant is predicted to change protein sequence (non-synonymous variants) or could affect expression. We also confirmed if any of the SNPs were possible regulatory elements or DNA features using RegulomeDB.^29^

DEPICT^30^ and FUMA^31^ were used to perform gene ontology and pathways analyses. We also leveraged brain single nuclei RNA expression data (http://ngi.pub/snuclRNA-seq/)^20^, to determine if the gene expression of the genes located in each identified loci was expressed in brain. For the ones expressed in brain, we also investigated if they were expressed in any specific brain cells (Figure 1B). Finally, we accessed blood RNA expression data taken at different times after stroke onset (3h, 5h and 24h) from the CLEAR trial^32^ (NCT00250991 at www.Clinical-Trials.gov) to test if the expression of genes located in the identified loci were associated with NIHSS or ΔNIHSS (NIHSS_5h_-NIHSS_24h_). We extracted the correlation between ΔNIHSS and gene expression (meausered using Affymetrix U133 Plus 2.0 array).^33^

### eQTL mapping, Mendelian Randomization and Colocalization

To identify the most likely functional gene, we accessed available expression quantitative trait (eQTL) datasets: the Genotype-Tissue Expression (GTEx) Project V8 (accessed on 10/10/2019), the Brain eQTL Almanac (Braineac) and an in-house dataset that includes brain expression data for 613 brains^34^. We used the summary-data-based Mendelian Randomization (SMR)^35^ and colocalization^36^ to test for pleiotropic association between the expression level of a gene and a complex trait to evaluate if the effect size of a genetic variant on the phenotype is mediated by gene expression (Figure 1B). We tested GWAS-significant and -suggestive loci from the ΔNIHSS analysis in two datasets: selected GTEx tissues (brain anterior cortex, cerebellum, brain cerebellar hemisphere, substantia nigra, hippocampus, frontal cortex, and putamen) and the Westra *et al*. dataset^37^ derived from whole blood. Both SMR and colocalization require effect sizes and the respective standard error to test the causal relationship, but MANTRA does not provide effect sizes. As a consequence, we used the summary statistics from the joint analysis for all populations to perform these analyses that are correlated with the results from MANTRA (r=-0.57; p<1.07×10^−05^ – data not shown). To complement the Mendelian randomization analyses with the posterior probability of a variant being causal in both GWAS and eQTL studies accounting for the genetic heterogeneity and *linkage disequilibrium* (LD), we used eCAVIAR^38^ which will consider several variants within the GWAS significant loci to perform the test.

### Genetic Correlation

We examined similarities in the genetic architectures of stroke early outcomes (ΔNIHSS) and stroke risk^15^ using PRSice^39^, LDSC^40^ and GNOVA^41^ (Figure 1B). Briefly, PRSice calculates polygenic risk scores at different p value thresholds by weighting each SNP by their effect size estimates. SNPs present in one dataset, ambiguous SNPs (A/T or C/G) and all SNPs in LD are removed prior to polygenic risk score calculation. LDSC and GNOVA estimate the genetic covariance and the variant-based heritability for two sets of summary statistics, each one corresponding to one trait of interest. These two parameters are used to calculate the genetic correlation and covariance respectively between the two traits. We limited our comparisons to the non-Hispanic White population to keep the population genetically homogeneous and use the 1000 Genomes European population-derived reference dataset. We calculated the genetic correlation between the European ischemic stroke summary statistics of the MEGASTROKE^15^ study and the non-Hispanic Whites meta-analysis summary statistics from the GENISIS study. We also determined if traits related to cardiovascular and general health (age at death^42^, lipid levels^43^ and body mass index (BMI)^44^) are genetically correlated to ΔNIHSS.

Summary statistics of the GENISIS dataset used for these analyses are available upon request. Individual data for the full GENISIS dataset will be uploaded to dbGAP titled: “*Genetics of Early Neurological Instability After Ischemic Stroke (GENISIS)”*.

## Results

The GENISIS study recruited 5,876 acute ischemic stroke patients from seven countries (Spain, Finland, Poland, United States, Costa Rica, Mexico and Korea). The mean patient age was 73 years; 45% of the patients were females; and 54% were treated with tPA. No significant differences in age or sex were found across sites. The distribution of TOAST classification of stroke etiology was also similar across sites. Significant differences were observed in baseline NIHSS and tPA treatment rates, likely due to differences in practices across the sites (Table 1).^6^ ΔNIHSS approximated a normal distribution, similar to that of each of the ethnic groups (non-Hispanic whites, Hispanics, African descent, and Asians) (Supplementary Figure 2).

**Table 1.** Demographic Characteristics of the GENISIS cohort by country.

|  | **Spain**<br>**(N=3,419)** | **Finland**<br>**(N=490)** | **Poland**<br>**(N=356)** | **US-EuA**<br>**(N=798)** | **Costa Rica**<br>**(N=141)** | **Mexico**<br>**(N=63)** | **Korea**<br>**(N=285)** | **US-AfA**<br>**(N=324)** | **GENISIS**<br>**(N=5,876)** |
| --- | --- | --- | --- | --- | --- | --- | --- | --- | --- |
| **Age** | 76.0 | 68.0 | 71.0 | 70.0 | 67.0 | 67.0 | 69.0 | 63.0 | 73.0 |
| **(years)\*** | (66.0-83.0) | (58.0-76.0) | (63.0-80.0) | (60.0-79.0) | (56.0-78.0) | (50.5-75.5) | (58.0-78.0) | (54.0-74.3) | (62.0-81.0) |
| **Sex** | 1,554 | 193 | 159 | 337 | 57 | 28 | 91 | 169 | 2,588 |
| **(Females, %)** | (45.5%) | (39.4%) | (44.7%) | (42.2%) | (40.4%) | (44.4%) | (31.9%) | (52.2%) | (44.0%) |
| **Baseline NIHSS\*** | 10.0 | 5.0 | 6.0 | 6.0 | 13.0 | 11.0 | 4.0 | 7.0 | 8.90 |
|  | (5.0-17.0) | (2.0-9.0) | (3.0-12.0) | (3.0-8.2) | (9.0-18.0) | (6.0-14.5) | (2.0-8.0) | (4.0-12.0) | (4.0-15.0) |
| **tPA Treatment (%)** | 48.32% | 48.37% | 59.55% | 73.81% | 100% | 46.03% | 28.07% | 75.62% | 54.20% |
| **ΔNIHSS** | 2.77±5.42 | 2.34±5.68 | 2.12±3.40 | 2.11±5.98 | 6.00±7.14 | 3.40±4.90 | 1.17±3.40 | 2.37±6.29 | 2.56±5.52 |
| **TOAST Classification\*\* (%):** |  |  |  |  |  |  |  |  |  |
| **Cardioembolic** | 38.32% | 41.63% | 29.21% | 37.72% | 21.28% | 23.81% | 30.53% | 29.01% | 36.50% |
| **Large Artery** | 17.17% | 16.53% | 12.36% | 13.03% | 39.01% | 25.40% | 24.56% | 8.64% | 16.76% |
| **Small Vessel Disease** | 9.15% | 6.73% | 3.09% | 13.16% | 12.77% | 14.29% | 17.89% | 16.98% | 10.14% |
| **Other** | 2.46% | 8.16% | 2.81% | 3.13% | 2.13% | 15.87% | 13.68% | 3.09% | 3.76% |
| **Undetermined** | 32.90% | 26.94% | 52.53% | 32.96% | 24.11% | 20.63% | 13.33% | 42.28% | 32.83% |

Values are expressed as mean±Standard Deviation. \*Values expressed as median (95% confidence interval). EuA: European American Ancestry, AfA: African American Ancestry.
\*\*TOAST classification criteria <sup>67</sup>

**Figure 2.**
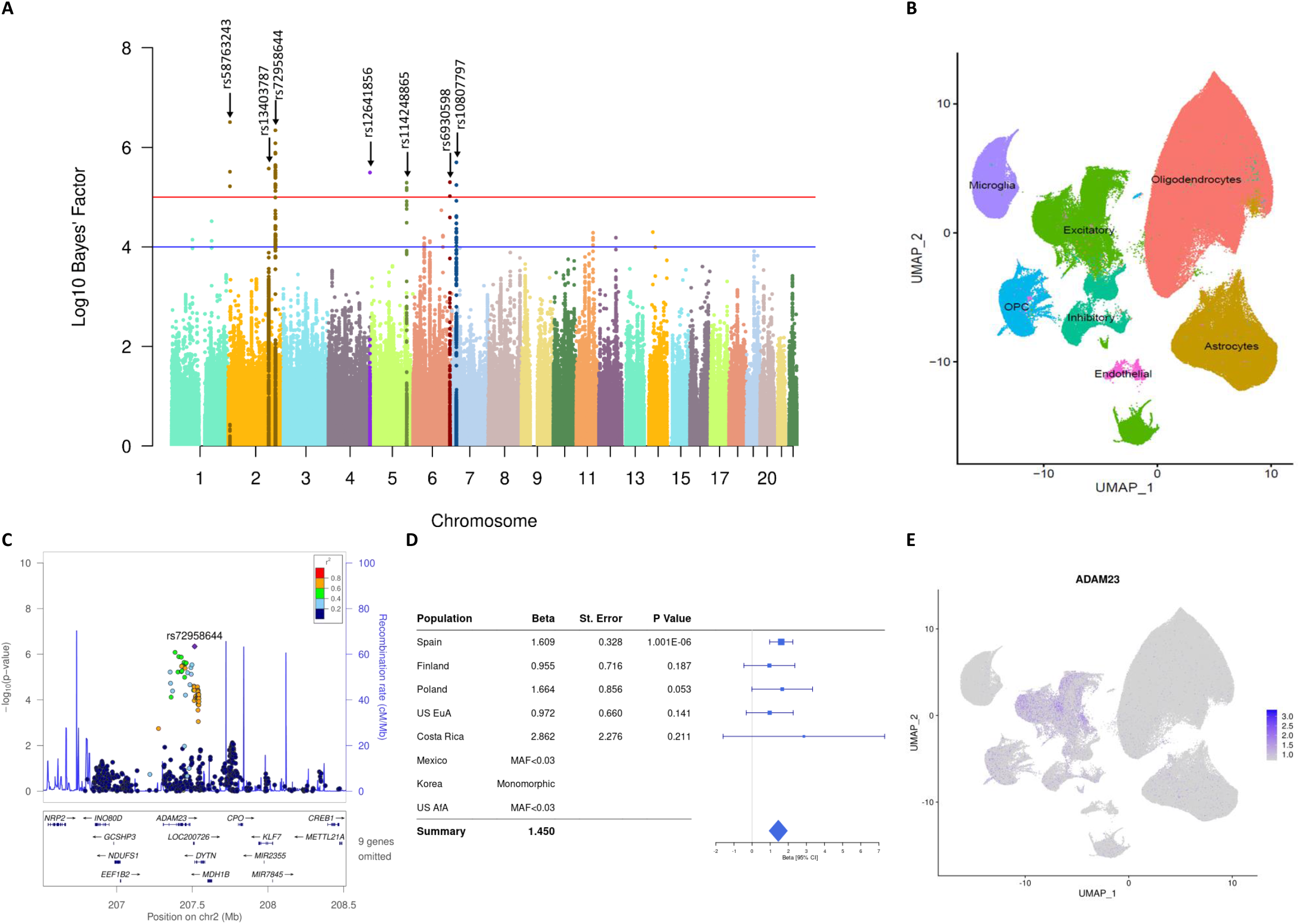
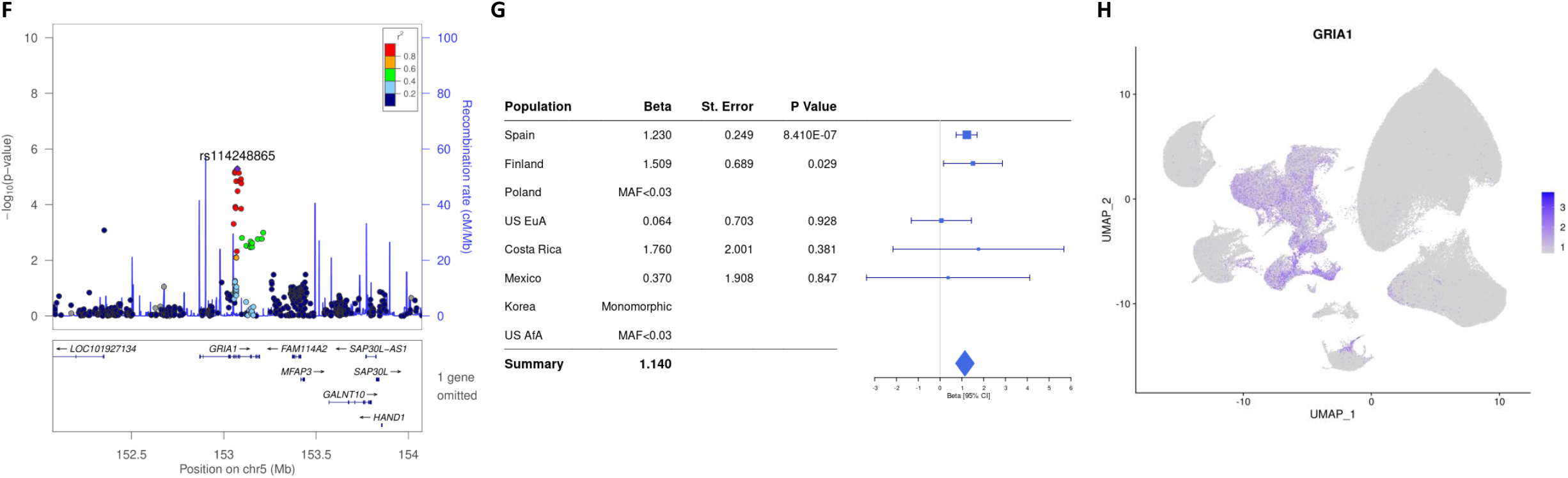
Association and annotation results. **A**. Association plot for ΔNIHSS. Manhattan plot shows LBF values from the multi-ancestry meta-analysis in each genomic location. The red line indicates the GWAS significant threshold (LBF>5) and the blue line the GWAS suggestive threshold (LBF>4). The genome-wide significant loci are highlighted. Local Manhattan plots are shown for **C**. rs72958644 and **F**. rs114248865 along with the corresponding forest plots, **D** and **G**, showing the contribution of each population to the overall signal. As part of the functional gene mapping, we accessed **B**. an in-house single nuclei data to describe the expression patterns in brain parietal lobe cell populations of the driving genes identified for **E**. rs72958644, *ADAM23* and **H**. rs114248865, *GRIA1*.

### Identification of novel loci associated with stroke early outcomes

We performed single variant analyses for each individual cohort separately; then we combined cohorts with similar ethnic backgrounds; finally, we performed a multi-ancestry meta-analysis with the four ethnic groups available in this study (Non-Hispanic Whites, Hispanics, Asians and African Americans) (Figure 1A). We identified seven GWAS significant loci (Figure 2A and Table 2) associated with ΔNIHSS.

**Table 2.**
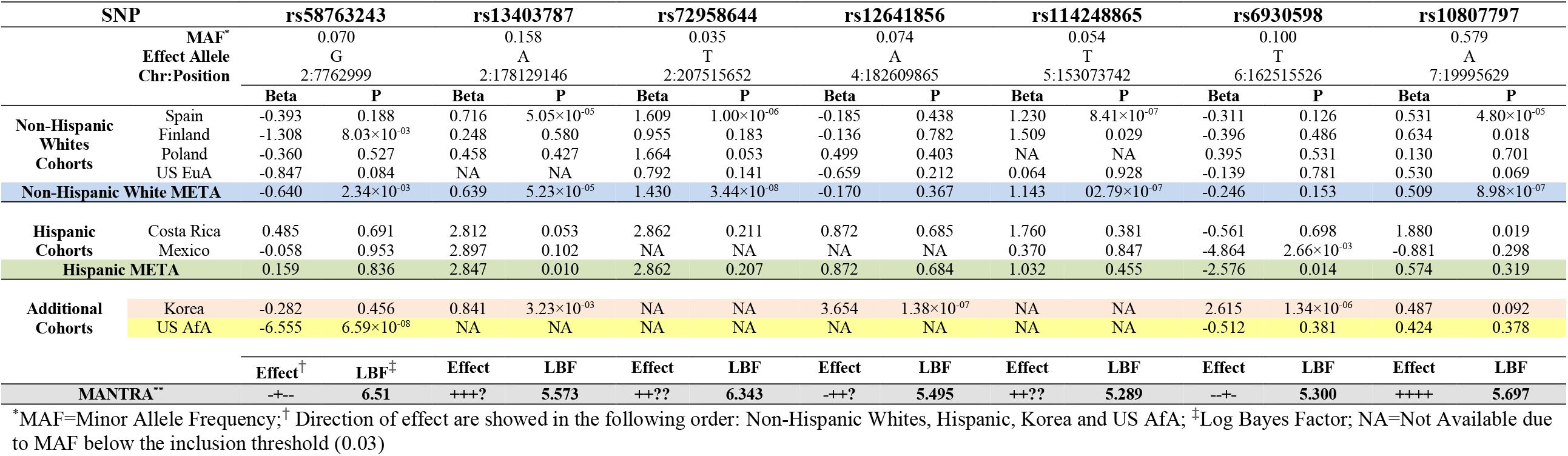
Summary Statistics for the Multi-Ancestry Meta-Analysis top hits by cohort.

Three independent loci were identified in chr2. The first locus, tagged by rs58763243 (MAF_G_=0.07; LBF=6.51), was located in a region comprised by several long non-coding RNAs and microRNAs (Supplementary Figure 3A). For this locus, all of the populations contributed to the association with negative betas, indicating that the minor allele was associated with lower (or more negative) ΔNIHSS. In addition this locus reached genome-wide significance in the US AfA population and was nominally significant in the Finnish population (Supplementary Figure 3B).

**Figure 3.**
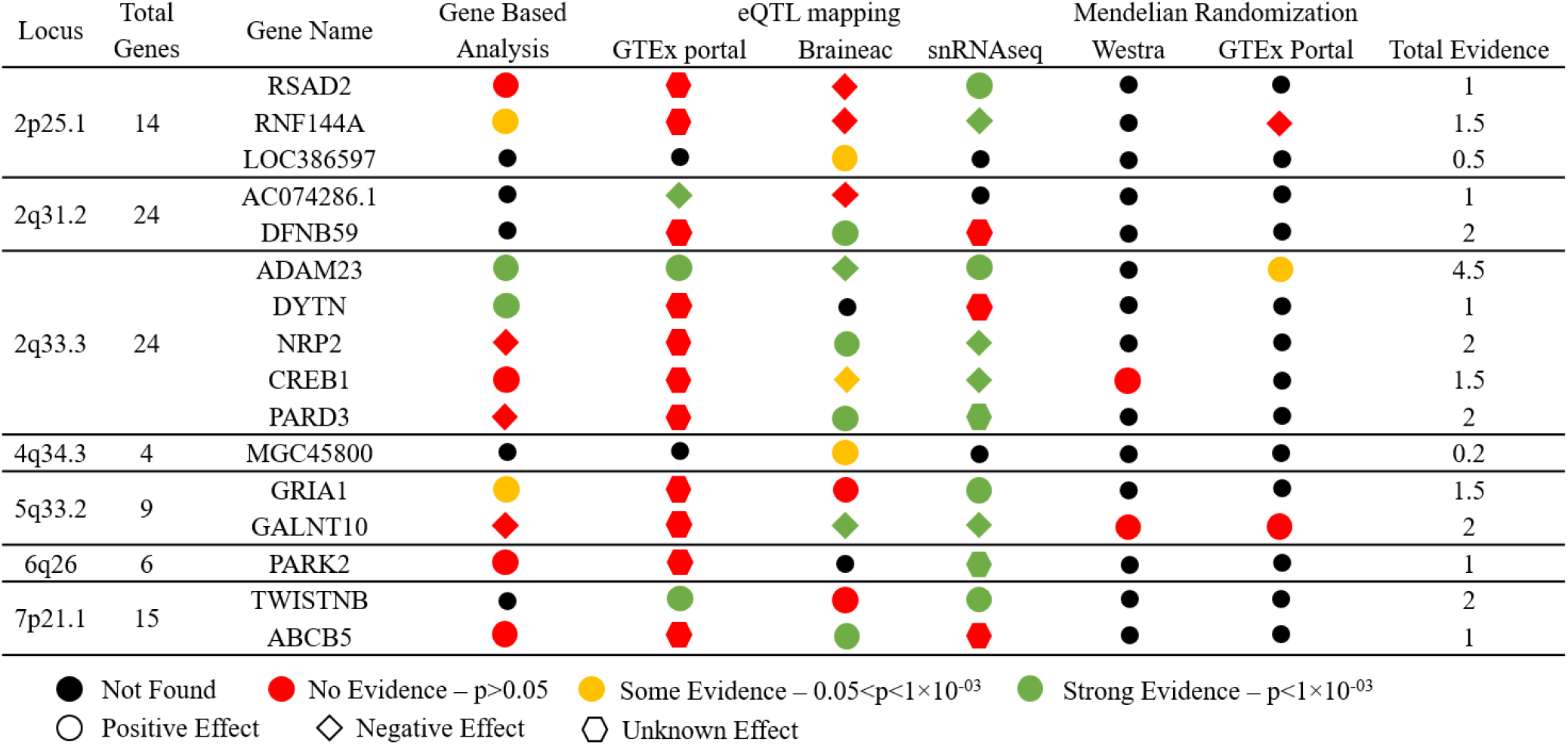
Gene prioritizing summary. Summary table showing the seven genome-wide significant loci from the multi-ancestry analysis (first column), the total number of genes identified in each of the locus (second column) and gene name for genes for which we have found some kind of evidence (third column). We have included the results from the gene-based analyses, the presence of any eQTL in GTEx portal or Braineac for any of the genome-wide or suggestive variants, if the gene is differentially expressed in neurons according to the snRNAseq data and the results from Mendelian randomization using Westra dataset (whole blood) or GTEx portal (all tissues). Black dots indicate that the gene was not found, red is that it was found but was not significant, yellow it was moderately significant (0.05<p<1×10^−03^) and green shows a significant association (p<1×10^−03^). Shape indicates the direction of effect, round positive, diamond negative and hexagon unknown.

The second locus, rs13403787 (MAF_A_=0.16; LBF=5.57), was also located on chr2 in a region with more than 20 genes between two recombination sites (Supplementary Figure 3C). The minor allele was associated with higher (or more positive) ΔNIHSS in all cohorts (Supplementary Figure 3D). The last genome wide significant locus in chr2 was rs72958644 (MAF_T_=0.04; LBF=6.34), located in a region that includes *ADAM23, CREB1, DYTN, NRP2, MDH1B*, among many others (Figure 2C). The signal is driven by the Non-Hispanic Whites (meta-analysis p=3.44×10^−08^), but virtually all ethnic groups contributed to this association, as the directionality was consistent across Hispanic, non-Hispanic White and AfA ethnic groups (Figure 2D and Supplementary Table 1). However, the SNPs in this locus were monomorphic in the Asian population. We observed a trend for positive correlation between ΔNIHSS with the genotype in this locus (R^2^=0.005, p=0.04; Supplementary Table 2).

Four additional loci were identified outside chr2. One locus at 4q34.3, rs12641856 (MAF_T_=0.05; LBF=5.50), in a region that includes four genes (*LINC00290, MGC45800, MIR1205* and *TENM3*; Supplementary Figure 3E). The signal was driven mainly by the Korean cohort (p=1.38×10^−07^) (Supplementary Figure 3F). The locus identified on chr5 is located on a region containing nine genes (*LOC101927134, GRIA1, FAM114A2, SAP30L, SAP30L-AS1, MFAP3, GALNT10, HAND1* and *MIR3141;* Figure 2F). The minor allele for the top hit in this locus, rs114248865 (MAF_T_=0.05; LBF=5.29) was associated with greater (more positive) ΔNIHSS across all cohorts, and was significant in the Spanish (p=8.41×10^−07^) and Finnish cohorts (p=0.03; Figure 2G). Another locus on chr6, tagged by rs6930598 (MAF_T_=0.06; LBF=5.30), was located in a region with the genes *PARK2, PACRG, MAP3K4, AGPAT4, AGPAT4-IT1* and *LOC101929239* (Supplementary Figure 3G). The variants in the region were significant in the Mexican and Korean cohorts but the direction of effect was not consistent (Supplementary Figure 3H). Moreover, the MAF for these variants ranged between 16% in the AfA population to 6% in the Asian and Finnish populations, suggesting that the region is very polymorphic depending on ethnicity. Thus, even though the locus is important for ΔNIHSS, it is possible that it is not the causal variant. Finally, we identified a locus on chr7, tagged by the variant rs10807797 (MAF_G_=0.34; LBF=5.70), and located in a gene rich region with 15 genes, including *TWISTNB, MACC1, TMEM196, ABCB5, RPL23P8* (Supplementary Figure 3I). This locus is tightly encompassed by two recombination sites. The top signal was significant or suggestive in all populations except the Polish and Mexican cohorts. Consistently, the direction of effect was the same in all cohorts except the Mexican cohort (Supplementary Figure 3J, Supplementary Table 1).

### Genetic contribution to early outcomes after ischemic stroke

We used GCTA to quantify the phenotypic variance explained by common SNPs. Because GCTA exploits LD patterns to calculate the explained variance, we restricted our analysis to non-Hispanic Whites. Due to founder effects present in the Finnish population, we also removed this cohort from the variance calculation (Final N=4,573). GTCA revealed that common genetic variants explained 8.7% of the variance of ΔNIHSS (p=0.001), confirming that genetic variants and genes are implicated on stroke outcomes. Next we determine what proportion of the genetic component is explained by the GWAS signals.

The SNPs comprised within the 7 genome-wide significant loci, defined as 500 bp upstream or downstream of the top signal, explained 2.1% of the total variance (p=1.80×10^−06^) of ΔNIHSS, or just 24.1% of the genetic component of ΔNIHSS. This suggests that there are additional loci associated with ΔNIHSS yet to be discovered. Thus, studies with larger sample size and more statistical power are needed to identify these additional loci.

### Functional Annotation of the Genome-Wide Significant Loci

Identifying the likely causal gene from each loci driving the association is a multi-step process (Figure 1B). We first annotated the suggestive variants (LBF>4), but none of them were predicted to change the protein sequence, comprise a regulatory element, or affect the chromatin architecture. Next, we explored publicly available datasets to investigate if any of the SNPs with suggestive LBFs were eQTLs (Supplementary Table 3). We performed gene-based analyses (Supplementary Table 4) and Mendelian Randomization (MR) analyses to identify possible causal relationships between gene expression and ΔNIHSS (Supplementary Table 5). Summary results can be found in Figure 3.

Gene-based analyses using FUMA suggested that *DYTN* (p=2.86×10^−05^, Z=4.02) and *ADAM23* (p=1.59×10^−04^, Z=3.60) were the genes driving the association at 2q33.3. Several variants in the *ADAM23* region were strong eQTLs for this gene in multiple tissues, based on the GTEx data (esophagus mucosa: p=1.90×10^−06^; Cultured Fibroblasts: p=3.70×10^−05^). Several SNPs in this region were also eQTLs for *NDFUS1* (thyroid p=1.20×10^−04^) although with less significant p-values than those for *ADAM23*. The variant was also significant in an independent dataset from the Braineac eQTL study, where rs13422013 (LFB=4.05) was also associated with *ADAM23* gene expression (thalamus p=7.60×10^−04^) and *NRP2* (cerebellum p=4.40×10^−04^). MR analyses indicated that *ADAM23* (p=0.05) and not *NRP2* (p>0.05) was the gene driving the association in this locus. Human brain single nuclei RNA-seq data indicate that *ADAM23* expression is enriched in neurons (p<2.20×10^−16^); compared to all the other brain cell types. More specifically, its expression is enriched in excitatory neurons (Figure 2E).

Gene-based analyses using FUMA revealed that *GRIA1* located in 5q33.2 was the gene most likely driving the association in that region (p=0.03, Z=1.83). However, Braineac identified several eQTLs for *GALNT10* (p=3.60×10^−04^) in the occipital cortex, but *GALNT10* did not reach statistical significance in the gene-based analysis. GTEx portal and the protein atlas reveals that *GRIA1* is mainly expressed in brain tissue. While *GALNT10* is also expressed in the brain, it has higher expression in other tissues. The human brain single nuclei RNA-seq data confirmed that both *GRIA1* and *GALNT10* are expressed in divergent brain cell types (Figures 2H and Supplementary 4A). *GRIA1* is highly-expressed in neurons compared to other cell types (p<2.20×10^−16^), but not expressed in oligodendrocytes (p<2.20×10^−16^) or astrocytes (p<2.20×10^−16^). In contrast, *GALNT10* is expressed in microglia, oligodendrocytes and astrocytes, but expression in neurons is low (p<2.20×10^−16^). *GRIA1* expression was also nominally associated with ΔNIHSS in the CLEAR trial dataset (p=0.002, r2=0.22).

Of the remaining five loci, we were able to map four (Supplementary Results). Briefly, eQTL analysis, revealed that 2q31.2 was likely to be driven by *DFNB59*, and 4q34.3 by *MGC45800*. No eQTLs were identified in 6q26, however, the locus falls within the boundaries of *PARK2*. Finally, 7p21.1 contains several eQTLs for *TWISTNB* and *ABCB5* (Supplementary Results).

### Pathway analyses

Gene ontology and pathway analyses using DEPICT and summary statistics for ΔNIHSS revealed consistent suggestive associations with functions relating to the brain and central nervous system. The top tissue enrichment from DEPICT identified the central nervous system (p=4.9×10^−03^), including the brain (p=4.9×10^−03^) and some brain regions: occipital lobe (p=1.52×10^−03^), parietal lobe (p=5.31×10^−03^), and hippocampus (p=5.07×10^−03^; Supplementary Table 6). The most significant pathways in the gene-set enrichment were the AKAP5 protein-protein interaction subnetwork (p=5.11×10^−04^), the CAMK1D protein-protein interaction subnetwork (p=5.27×10^−04^), the RALA protein-protein interaction subnetwork (p=6.32×10^−04^) and neurotransmitter uptake (p=6.74×10^−04^; Supplementary Table 7). Several genome-wide significant candidate genes fell within these networks, including *AGPS* (2q31.2, LBF=5.57) in the RALA subnetwork and *GPRS1* (2q33.3, LBF=6.34) in the CAMK1D subnetwork. AKAP5, expressed almost uniquely in brain (GTEx portal and the protein atlas.), interacts with post-synaptic density protein 95 (PSD-95) in dendritic spines and modulates synaptic transmission.^45-47^ Both CAMK1D and RALA, expressed in most tissues (CAMK1D demonstrates higher expression in brain), are involved in signal transduction functions. MAGMA gene-set analyses revealed the gene-set “*meissner_brain_hcp_with_h3k4me3_and_h3k27me3*” (p=9.41×10^−06^, Supplementary Table 8). Unfortunately, there is no function associated with this gene set that has been described in animal models. Together these results indicate that our unbiased genetic screening has identified genes that are largely expressed in the brain and in some cases involved in pathways associated with neuronal and synaptic function.

### Unique genetic architecture of early outcomes after stroke

We examined the genetic architecture of ΔNIHSS for shared genetic variation and variance with other cardiovascular and aging-related traits, including stroke risk, age at death, plasma lipid levels and body mass index using PRSice (Supplementary Table 9 and Figure 5), LDSC (Supplementary Table 10) and GNOVA (Supplementary Table 11). Supported by a previous report^18^, PRSice did not reveal any genetic overlap between the genetic architecture of ΔNIHSS and stroke risk. Similarly, no overlap with age at death, lipid levels, or BMI was found. LDSC was unable to calculate the heritability estimate for ΔNIHSS. GNOVA, was successful at estimating the heritability for ΔNIHSS, and revealed significant covariance with stroke risk, but only when correcting for sample overlap (p=4.67×10^−05^, correlation=0.80) and age at death (p=9.26×10^−04^, correlation=1). However, the heritability estimate for ΔNIHSS for overlap with lipid levels and body mass index was negative, likely due to the low number of variants included in the analyses. Because both GNOVA and LDSC require larger sample sizes, the results of these analyses were inconclusive.

## Discussion

The first 24 hours after stroke onset is a period of great neurological instability, which may reflect brain tissue at risk for infarction but with the potential for salvageability.^4,48-50^ Not only is early neurological change common, it is influenced by known mechanisms involved in early deterioration/improvement, and has a strong influence on long-term functional outcome.^6^ Here, we performed a GWAS using ΔNIHSS as a quantitative phenotype in 5,876 acute ischemic stroke patients. We found that ΔNIHSS is heritable: common SNPs account for 8.7% of its variance. We have found seven genome-wide significant loci that are related to ΔNIHSS. However, they explain only 2.1% of the variance, indicating that 6.6% of the variance is explained by genes below the genome-wide significant threshold. Through functional annotation, we have linked each locus to specific genes, some of which are uniquely expressed in the brain.

Of all the loci showing association with ΔNIHSS, functional annotation analyses strongly suggest functional genes for two of the seven loci: *ADAM23* for 2q33.3 and *GRIA1* for 5q33.2. ADAM23 belongs to the ADAM (a disintegrin and metalloproteinase) family of proteins, defined by a single-pass transmembrane structure with a metallopeptidase domain (some inactive). This protein family is involved in cell adhesion, migration, proteolysis and signaling.^51^ ADAM23 is a transmembrane member without catalytic domain, and is involved in cell-cell and cell-matrix interactions.^51,52^ Previous studies have shown that *ADAM23* is expressed in pre-synaptic membranes, linked by the extracellular protein LGI1 to post-synaptic ADAM22.^53,54^ We found that *ADAM2*3 was expressed primarily in excitatory neurons of the cerebral cortex, based on our human brain single-nuclei transcriptomics dataset^20^, and confirmed by the Human Transcriptomic Cell Types dataset from the Allen Brain Map.^55^ Several lines of evidence suggest that ADAM23 is important for pathological synaptic excitability: 1) *adam23* is a common risk gene for canine idiopathic epilepsy;^56-58^ 2) mutations in its binding partner, *LGI1*, cause the neurological syndrome, ADPEAF (autosomal dominant partial epilepsy with auditory features)^59^; 3) autoimmunity against LGI1 (as seen in limbic encephalitis) results in seizures and encephalopathy.^60^

Indeed, ADAM23 is also known to be a binding partner (via ADAM22 and PSD95) of the protein product of another one of our genome-wide significant associated genes, *GRIA1*, which encodes for the α-amino-3-hydroxy-5-methyl-4-isoxazolepropionic acid receptor subunit 1 (AMPAR1).^54^ It has long been known that AMPA receptors, along with other glutamate receptors, are mediators of excitotoxic neuronal death, hypothesized to play an important role in ischemic brain injury.^61,62^ The failure of numerous older clinical trials examining the efficacy of anti-excitotoxic drugs has cast doubt on the relevance of excitotoxicity in human acute ischemic stroke, although questions about the quality of these early clinical trials have been raised.^63-66^ Thus, the association between *ADAM23* and *GRIA1* with ΔNIHSS provides the first genetic evidence that excitotoxicity may contribute to ischemic brain injury in humans.

The plausible roles that *ADAM23* and *GRIA1* play in acute brain ischemia mechanisms lend support to the idea that GWAS using ΔNIHSS as a quantitative phenotype can identify novel mechanisms and potential drug targets to mitigate neurological deterioration or enhance early improvement after stroke. From the CLEAR dataset, ^32,33^ expression levels of *GRIA1* in peripheral blood of ischemic stroke patients was associated with ΔNIHSS between 5h and 24h post stroke onset, supporting a link between increased expression of GRIA1 and improved outcomes. In addition to the two genes discussed above, our GWAS identified five other loci—the functional genes remain to be identified. Acute ischemic stroke patients are extremely well-phenotyped, as part of standard of care, with both clinical assessments and structural/physiological imaging. Thus, there is great potential for additional quantitative phenotypes to expand understanding of the genetic architecture of acute ischemic stroke, promising to identify novel mechanisms and drug targets. Larger and more comprehensive genetic studies of acute ischemic stroke are needed.

There are several limitations to this study. GENISIS enrolled a heterogeneous group of stroke patients without regard to underlying etiology, stroke localization and genetic and environmental background. Although we have previously demonstrated that etiology (TOAST criteria) has little influence on ΔNIHSS, it is likely that mechanisms involved in neurological instability may depend on etiology. Stroke localization may also be an important determinant of mechanisms involved in neurological instability. For example, mechanisms in cortical strokes may differ from those in subcortical or brainstem strokes. False positive findings due to the characteristics of the population is possible, but by using MANTRA we were able to correct by population heterogeneity. Future studies might aim to enroll a more homogeneous cohort of stroke patients to increase power to discover more genetic variants that associate with neurological instability. Finally, most of the patients in GENISIS were enrolled prior to the thrombectomy treatment era, and patients that underwent thrombectomy were excluded from the study to reduce heterogeneity. As a result, genetic interactions with reperfusion are largely unexplored.

## Summary

In conclusion, we identified seven novel loci associated with early neurological instability after ischemic stroke. Two of these loci were linked to genes, *ADAM23* and *GRIA1*, involved in synaptic excitability at glutamatergic synapses.

These results provide some of the first evidence that excitotoxic mechanisms are relevant to acute ischemic stroke in humans. Moreover, these results provide proof of principle that GWAS using ΔNIHSS, a quantitative phenotype of early neurological instability, may reveal underlying mechanisms involved in ischemic brain injury.

## Supporting information

Supplementary Methods and Results

## Data Availability

Summary statistics of the GENISIS dataset used for these analyses are available upon request. Individual data for the full GENISIS dataset will be uploaded to dbGAP titled: Genetics of Early Neurological Instability After Ischemic Stroke (GENISIS).

## Acknowledgments

We would like to thank the patients and their families for making possible all the genetic studies included in this manuscript. We also thank the MEGASTROKE consortium for access to the data (see full list of MEGASTROKE authors in supplementary data), the Genotype-Tissue Expression (GTEx) Project (supported by the Common Fund of the Office of the Director of the National Institutes of Health, and by NCI, NHGRI, NHLBI, NIDA, NIMH, and NINDS) and the Brain eQTL Almanac (Braineac) resource to access the UK Brain Expression Consortium (UKBEC) dataset.

