## Supplementary Figures for "Multi-ancestry genetic study in 5,876 patients identifies an association between excitotoxic genes and early outcomes after acute ischemic stroke"

**Supplementary Figure 1. Correlation between models.** We tested the association of SNPs across the genome with ΔNIHSS using an additive linear model adjusted by sex, age, the two Principal Components SNP genotyping batch, TOAST classification and baseline NIHSS. To test the effect of tPA on the results, we perform a joint analysis in the complete GENISIS cohort including tPA as covariate and compare it to the same model without tPA. The figure show how in both models, the results are highly correlated. The X-axis shows the negative $log_{10}$-transformed p-values for the model without tPA; the Y-axis shows the negative $log_{10}$-transformed p-values for the model with tPA as covariate

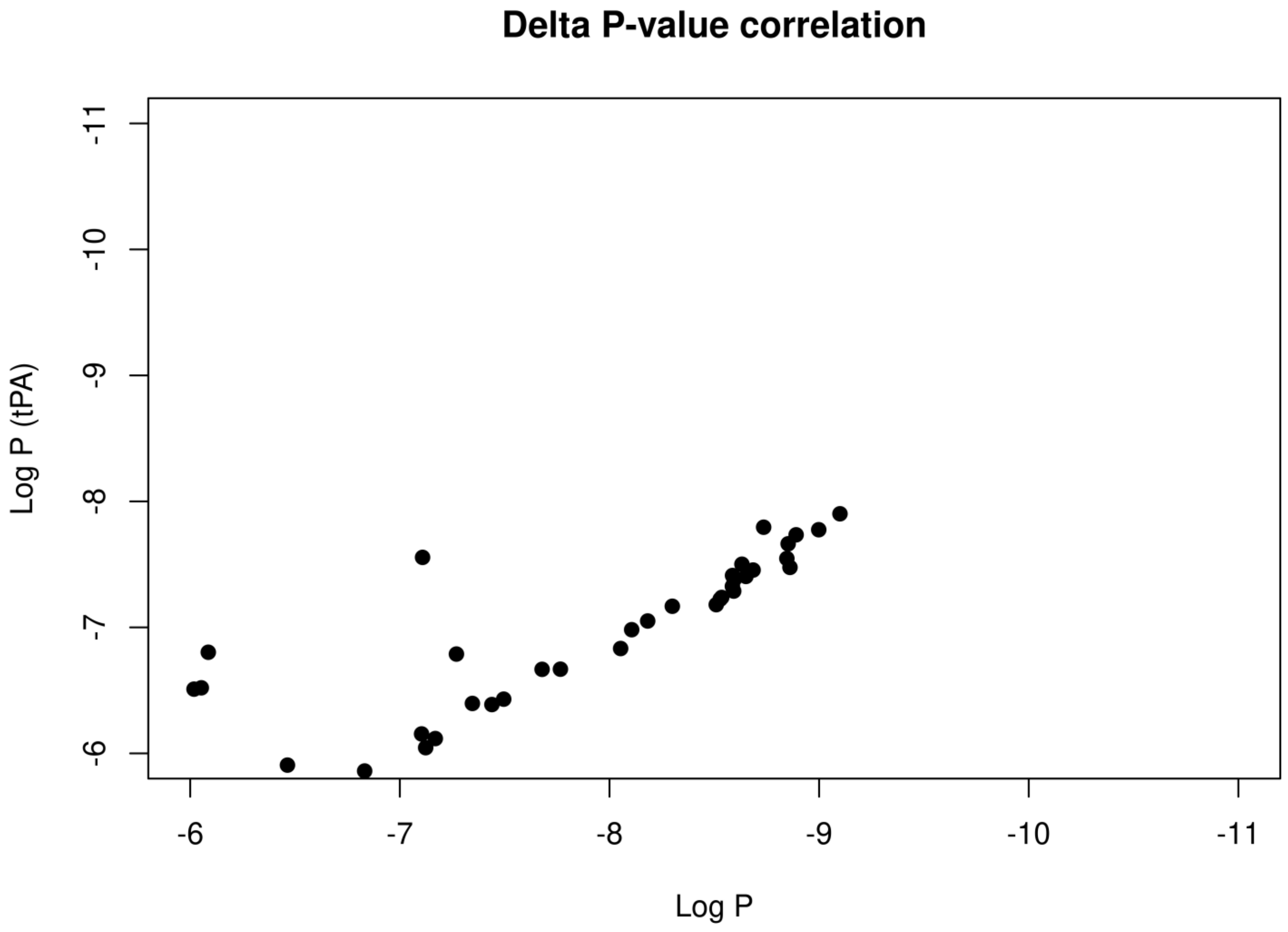

**Supplementary Figure 2. ΔNIHSS Distribution. A.** Distribution of ΔNIHSS (blue), baseline NIHSS (green) and 24h NIHSS (red) in the complete GENISIS cohort. **B.** Distribution of ΔNIHSS by ethnic background; non-Hispanic white (blue), Hispanic (green), Korea (orange) and US participants with African descent (US AfA – yellow).

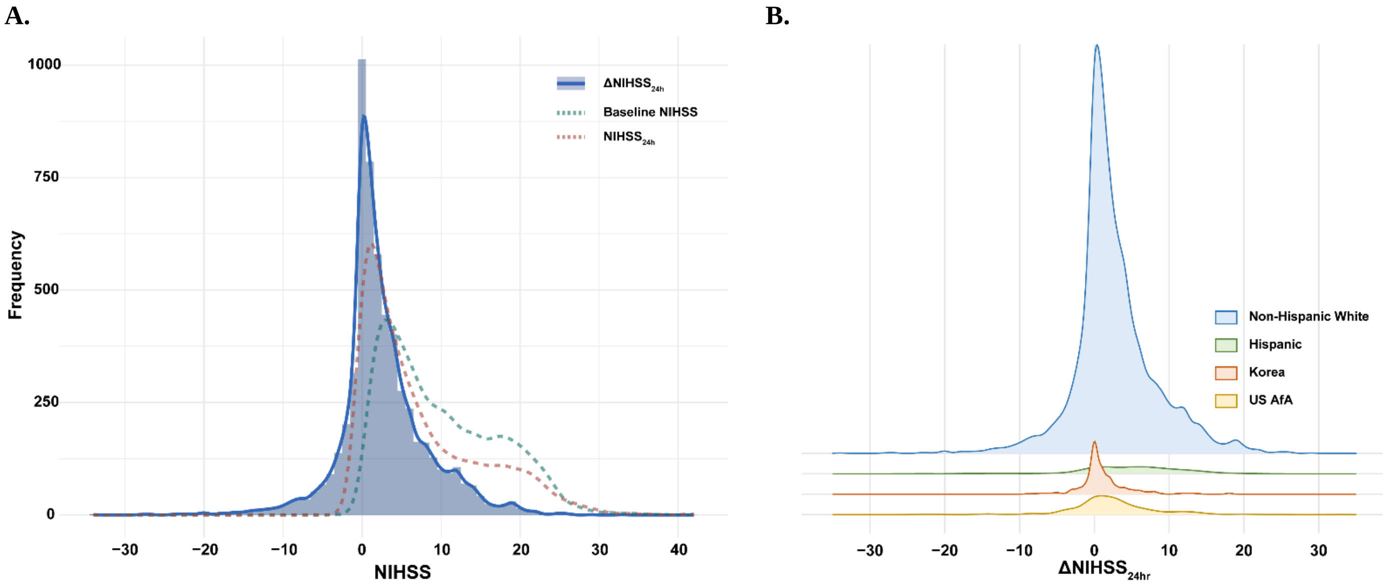

**Supplementary Figure 3. Association results.** Local Manhattan plots for **A.** rs58763243, **C.** rs13403787, **E.** rs12641856, **G.** rs6930598 and **I.** rs10807797 along with the corresponding forest plots, **B**, **D**, **F**, **H** and **J**, showing the contribution of each population to the overall signal.

**A.**

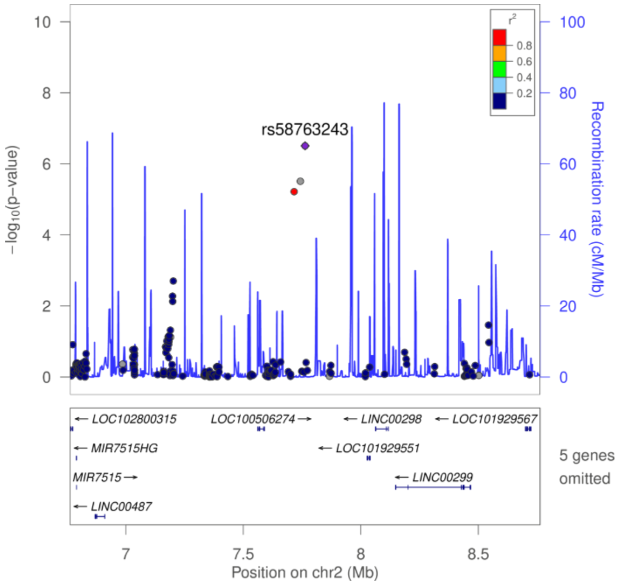


**B.**

| Population | Beta | St. Error | P Value |
|---|---|---|---|
| Spain | -0.393 | 0.298 | 0.188 |
| Finland | -1.308 | 0.491 | 8.028E-03 |
| Poland | -0.360 | 0.569 | 0.527 |
| US EuA | -0.847 | 0.490 | 0.084 |
| Costa Rica | 0.485 | 1.217 | 0.953 |
| Mexico | -0.058 | 0.994 | 0.691 |
| Korea | -0.282 | 0.378 | 0.426 |
| US AfA | -6.555 | 1.183 | 6.585E-08 |
| **Summary** | **0.720** | | |

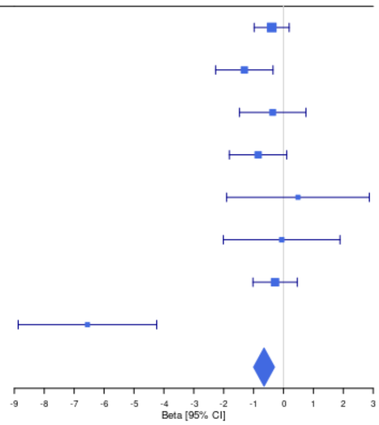


**C.**

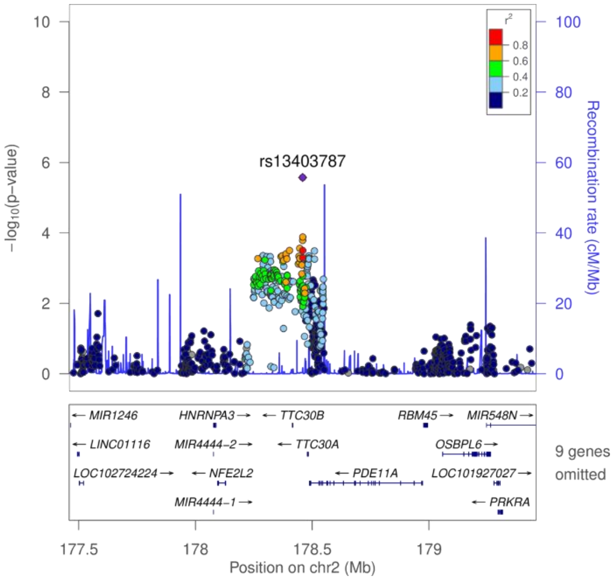


**D.**

| Population | Beta | St. Error | P Value |
|---|---|---|---|
| Spain | 0.716 | 0.176 | 5.054E-05 |
| Finland | 0.248 | 0.448 | 0.580 |
| Poland | 0.458 | 0.575 | 0.427 |
| US EuA | | | |
| Costa Rica | 2.812 | 1.438 | 0.053 |
| Mexico | 2.897 | 1.738 | 0.102 |
| Korea | 0.841 | 0.284 | 0.003 |
| US AfA | | | |
| **Summary** | **0.72** | | |

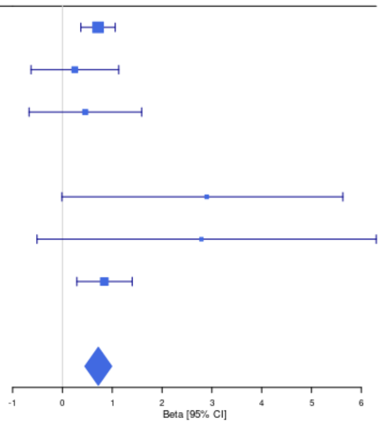

**E.**

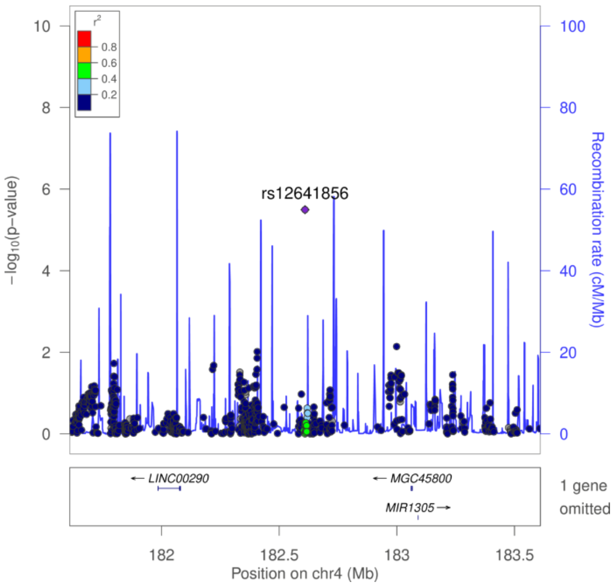


**F.**

| Population | Beta | St. Error | P Value |
|---|---|---|---|
| Spain | -0.185 | 0.239 | 0.438 |
| Finland | -0.136 | 0.491 | 0.782 |
| Poland | 0.499 | 0.596 | 0.403 |
| US EuA | -0.659 | 0.528 | 0.212 |
| Costa Rica | 0.872 | 2.144 | 0.685 |
| Mexico | MAF<0.03 | | |
| Korea | 3.715 | 0.686 | 1.378E-07 |
| US AfA | MAF<0.03 | | |
| **Summary** | **0.110** | | |

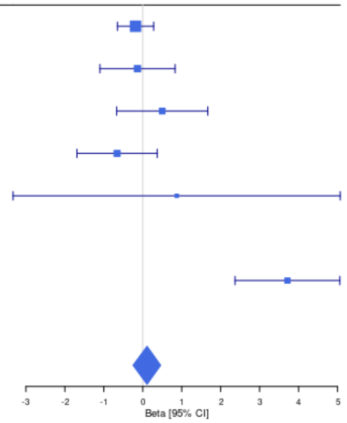


**G.**

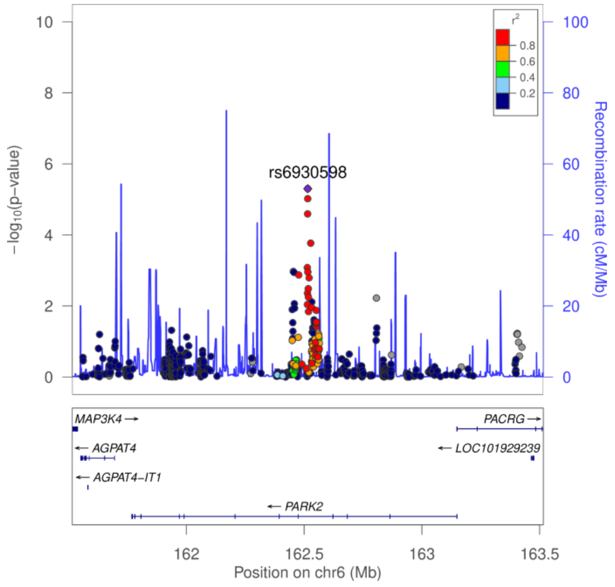


**H.**

| Population | Beta | St. Error | P Value |
|---|---|---|---|
| Spain | -0.311 | 0.203 | 0.126 |
| Finland | -0.396 | 0.568 | 0.486 |
| Poland | 0.395 | 0.629 | 0.531 |
| US EuA | -0.139 | 0.498 | 0.781 |
| Costa Rica | -0.561 | 1.444 | 0.698 |
| Mexico | -4.864 | 1.539 | 0.003 |
| Korea | 2.615 | 0.529 | 1.33E-06 |
| US AfA | -0.512 | 0.583 | 0.381 |
| **Summary** | **-0.070** | | |

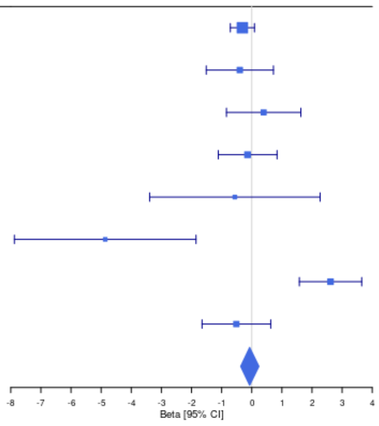


**I.**

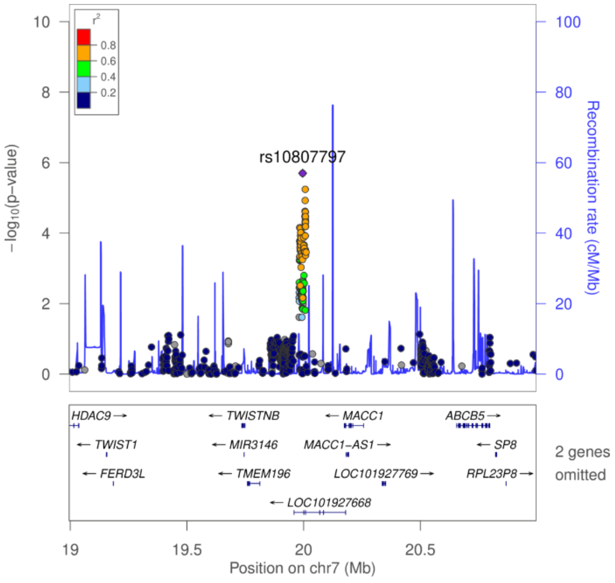


**J.**

| Population | Beta | St. Error | P Value |
|---|---|---|---|
| Spain | -0.531 | 0.131 | 4.804E-05 |
| Finland | -0.634 | 0.268 | 0.018 |
| Poland | -0.130 | 0.337 | 0.701 |
| US EuA | -0.530 | 0.291 | 0.069 |
| Costa Rica | -1.880 | 0.793 | 0.019 |
| Mexico | 0.881 | 0.838 | 0.298 |
| Korea | 0.487 | 0.288 | 0.092 |
| US AfA | 0.425 | 0.481 | 0.378 |
| **Summary** | **-0.370** | | |

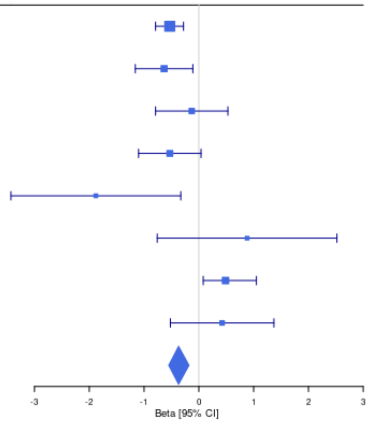

**Supplementary Figure 4. Single-nuclei data on brain cortex.** Expression patterns across the brain cortex cell populations derived from single nuclei RNAseq data for  for **A.** *GALNT10*, candidate gene along with *GRIA1* for the genome wide locus in chr.5 lead by rs114248865; **B.** *DFNB59* also known as *PJVK*, candidate gene for the genome wide locus in chr.2 lead by rs13403787, **C.** *TWISTNB*, candidate gene for the genome wide locus in chr6 along with **D.** *ABCB5* lead by rs10807797.

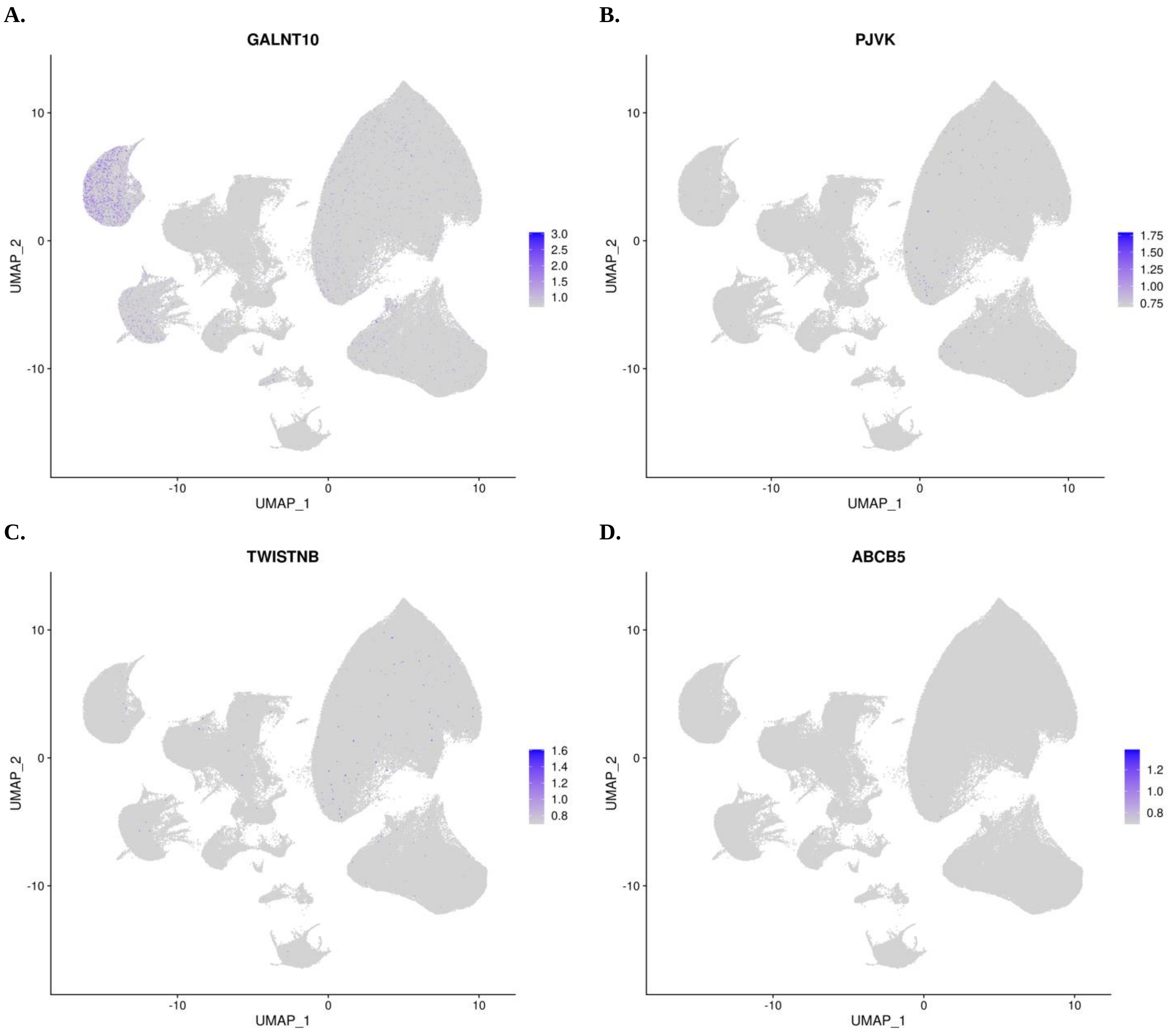

**Supplementary Figure 5. PRSice results.** We used the software PRSice2 to perform comparisons using polygenic risk scores at different p-value thresholds between the genetic architecture of ΔNIHSS and **A and B.** stroke risk, **C and D** age at death, **E and F** total cholesterol, **G and H** triglycerides, **I and J** high density lipoproteins, **K and L** low density lipoproteins and **M and N** body mass index.

**A.**

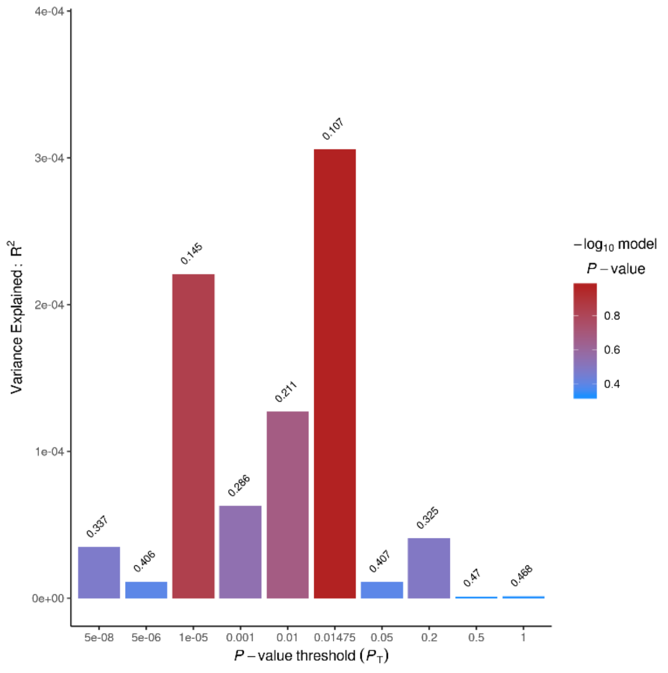


**B.**

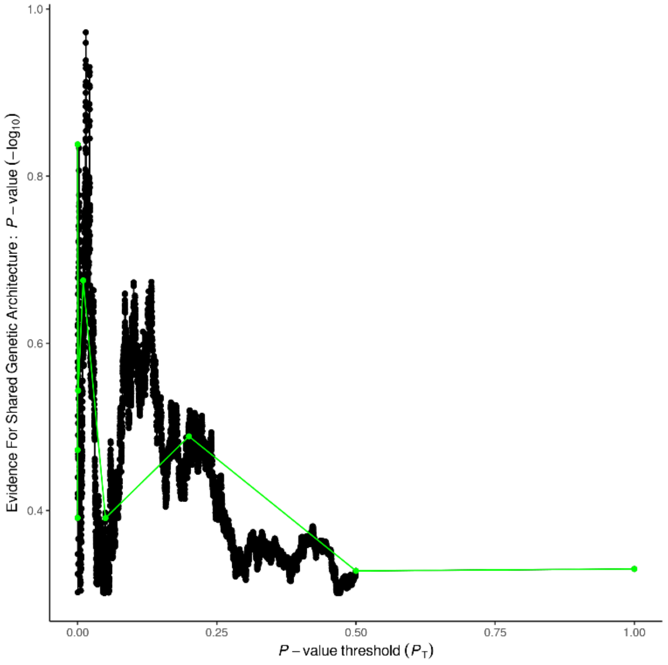


**C.**

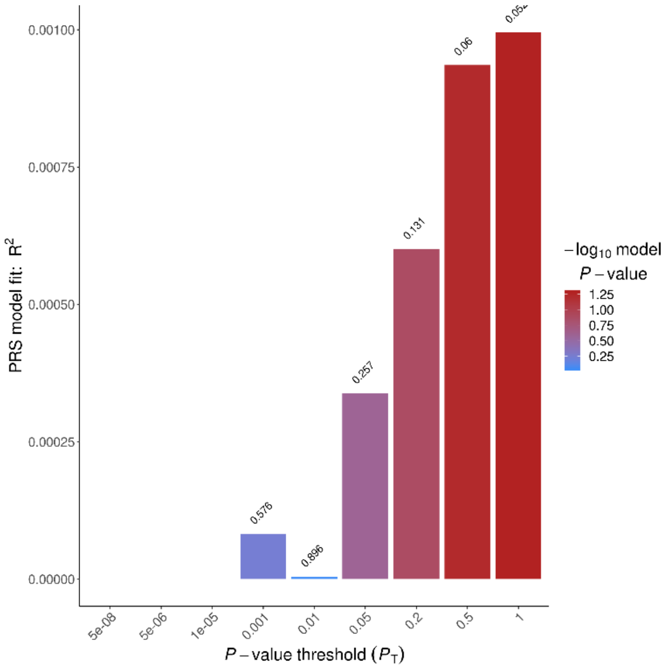


**D.**

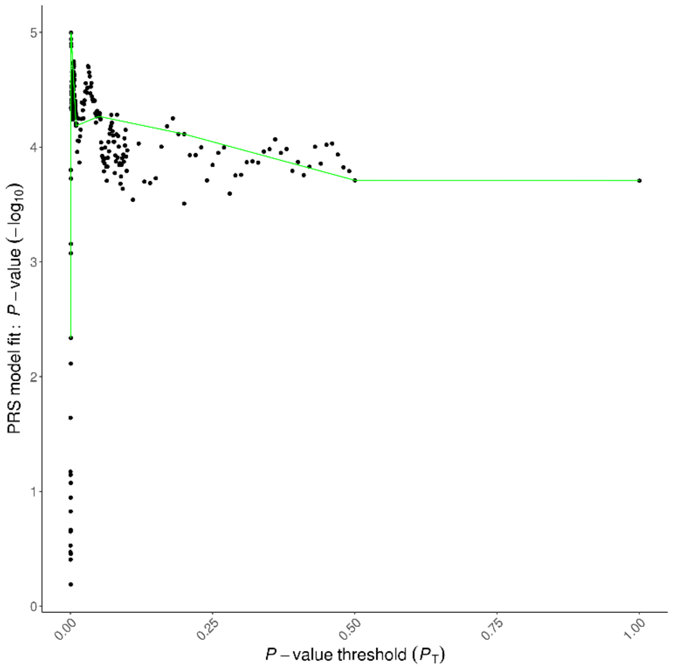

E.

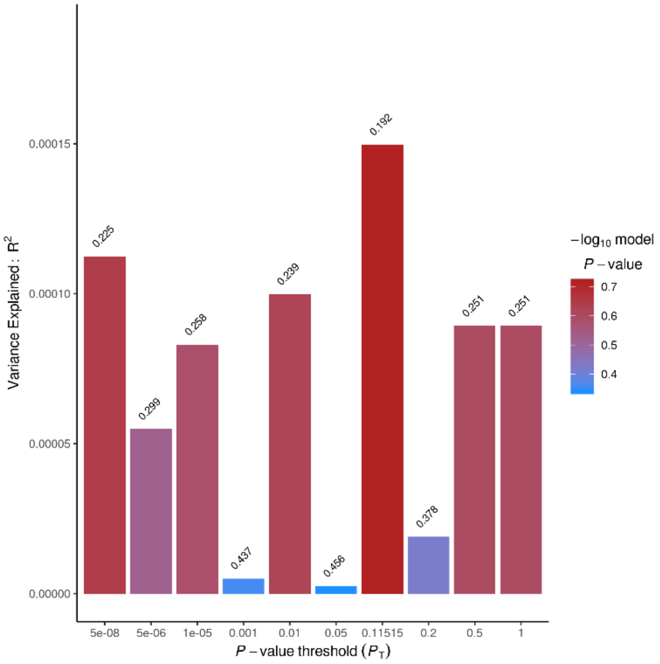

Variance Explained : R2
0.00015
0.00010
0.00005
0.00000
0.225
0.299
0.258
0.437
0.239
0.456
0.192
0.378
0.251
0.251
5e-08
5e-06
1e-05
0.001
0.01
0.05
0.11515
0.2
0.5
1
P − value threshold (PT)
−log10 model
P − value
0.7
0.6
0.5
0.4


F.

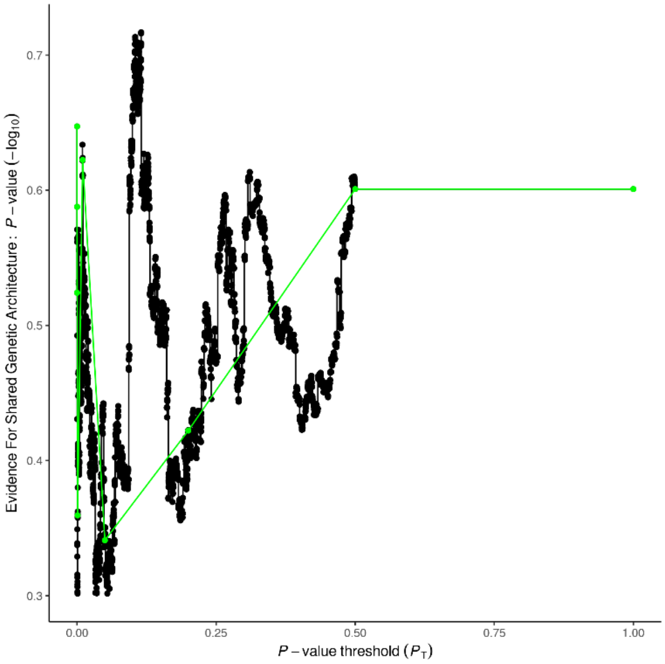

Evidence For Shared Genetic Architecture : P − value (−log10)
0.7
0.6
0.5
0.4
0.3
0.00
0.25
0.50
0.75
1.00
P − value threshold (PT)


G.

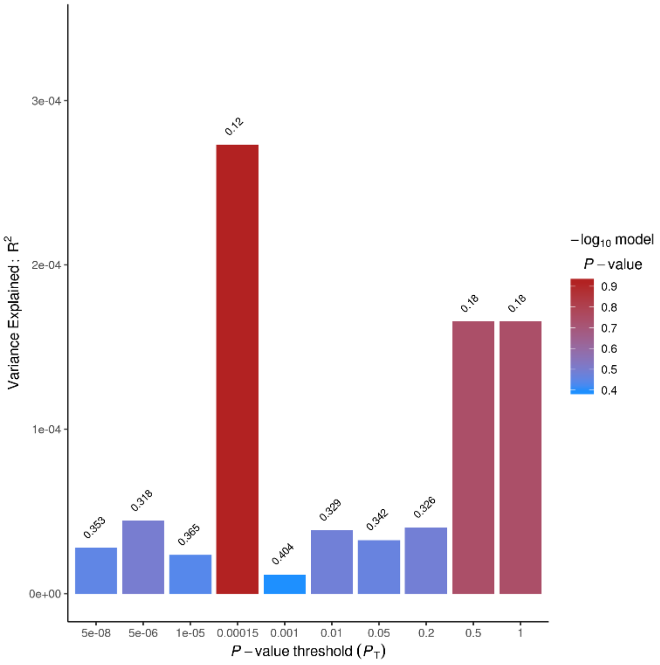

Variance Explained : R2
3e-04
2e-04
1e-04
0e+00
0.353
0.318
0.365
0.12
0.404
0.329
0.342
0.326
0.18
0.18
5e-08
5e-06
1e-05
0.00015
0.001
0.01
0.05
0.2
0.5
1
P − value threshold (PT)
−log10 model
P − value
0.9
0.8
0.7
0.6
0.5
0.4


H.

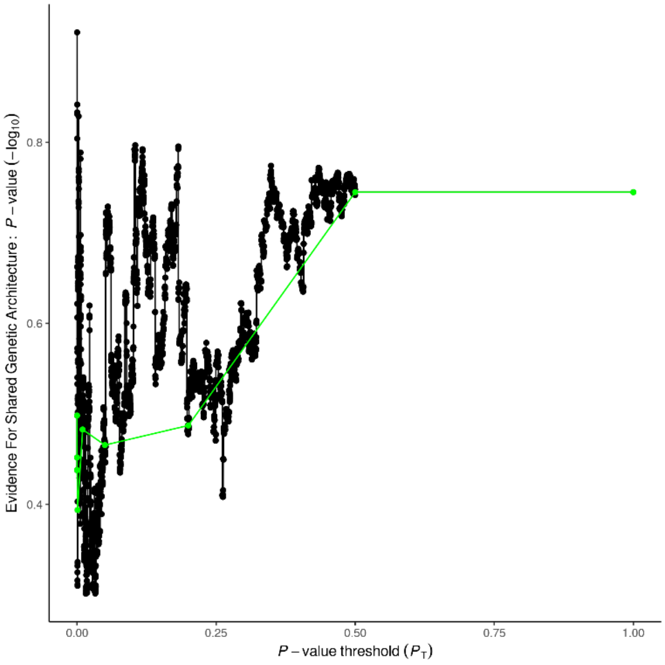

Evidence For Shared Genetic Architecture : P − value (−log10)
0.8
0.6
0.4
0.00
0.25
0.50
0.75
1.00
P − value threshold (PT)

**I.**

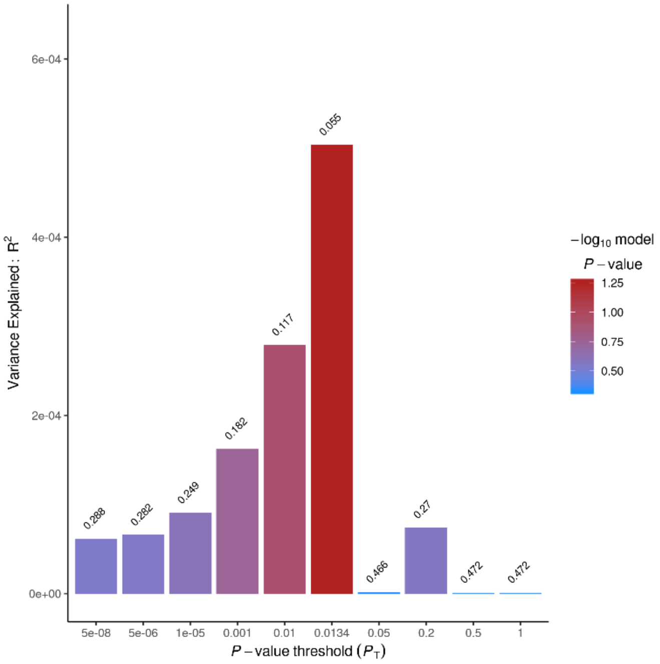
Variance Explained : $R^2$
6e-04
4e-04
2e-04
0e+00
0.288
0.282
0.249
0.182
0.117
0.055
0.466
0.27
0.472
0.472
5e-08
5e-06
1e-05
0.001
0.01
0.0134
0.05
0.2
0.5
1
P – value threshold ($P_T$)
$-\log_{10}$ model P – value
1.25
1.00
0.75
0.50

**J.**

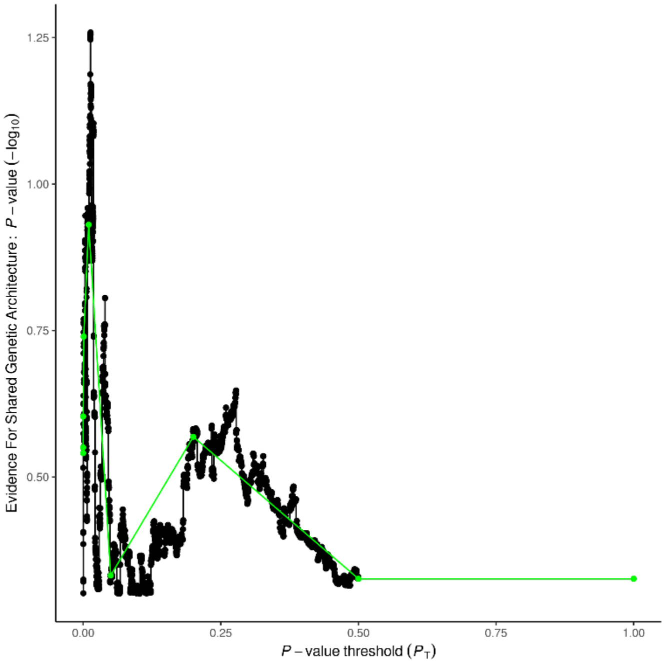
Evidence For Shared Genetic Architecture : P – value ($-\log_{10}$)
1.25
1.00
0.75
0.50
0.00
0.25
0.50
0.75
1.00
P – value threshold ($P_T$)

**K.**

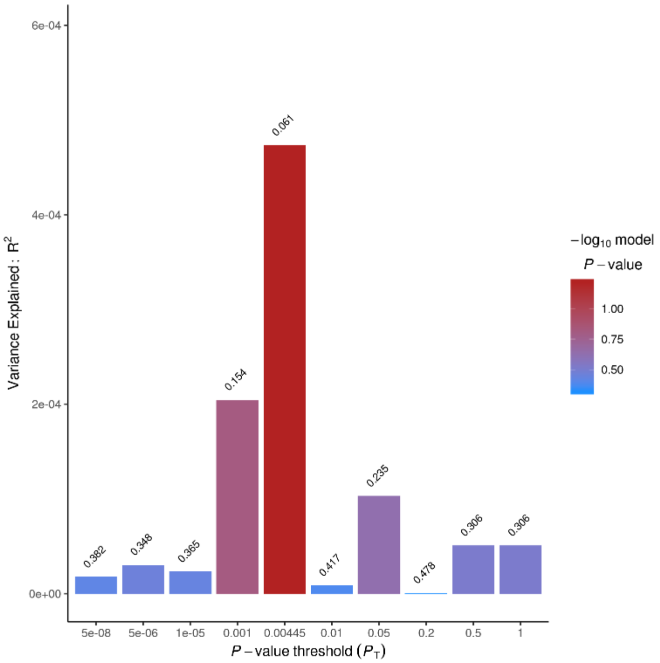
Variance Explained : $R^2$
6e-04
4e-04
2e-04
0e+00
0.382
0.348
0.365
0.154
0.061
0.417
0.235
0.478
0.306
0.306
5e-08
5e-06
1e-05
0.001
0.00445
0.01
0.05
0.2
0.5
1
P – value threshold ($P_T$)
$-\log_{10}$ model P – value
1.00
0.75
0.50

**L.**

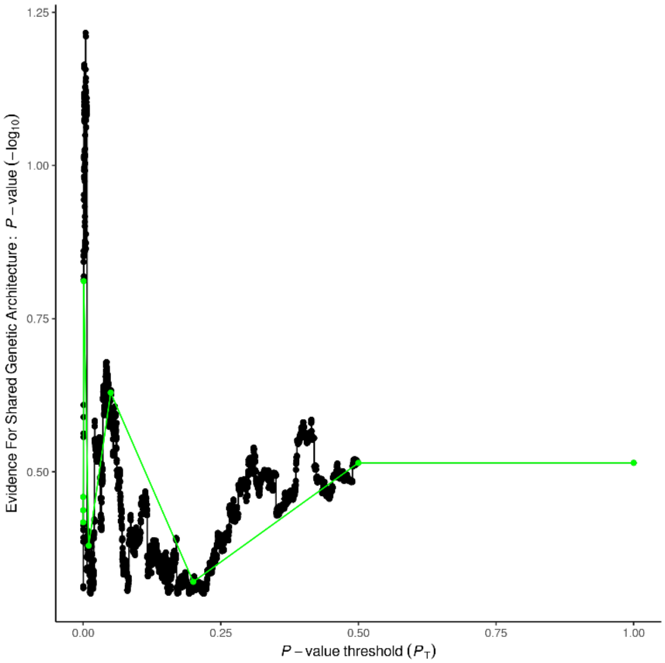
Evidence For Shared Genetic Architecture : P – value ($-\log_{10}$)
1.25
1.00
0.75
0.50
0.00
0.25
0.50
0.75
1.00
P – value threshold ($P_T$)

**M.**

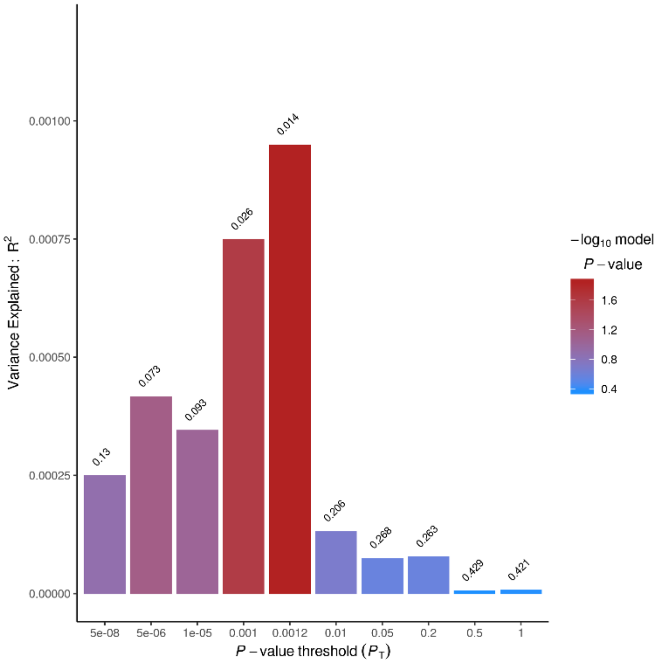
Variance Explained : $R^2$
0.00100
0.00075
0.00050
0.00025
0.00000
0.13
0.073
0.093
0.026
0.014
0.206
0.268
0.263
0.429
0.421
5e-08
5e-06
1e-05
0.001
0.0012
0.01
0.05
0.2
0.5
1
$P$ − value threshold ($P_T$)
$-\log_{10}$ model $P$ − value
1.6
1.2
0.8
0.4

**N.**

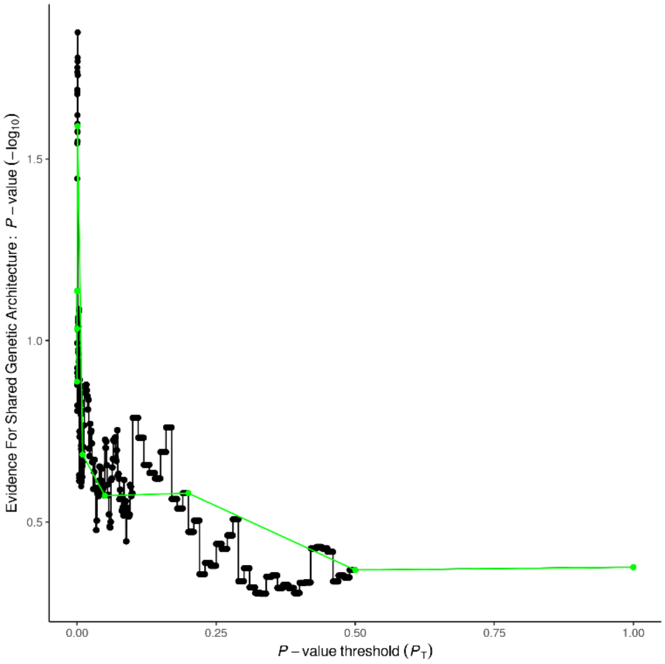
Evidence For Shared Genetic Architecture : $P$ − value ($-\log_{10}$)
1.5
1.0
0.5
0.00
0.25
0.50
0.75
1.00
$P$ − value threshold ($P_T$)
