## Supplementary Methods and Results for "Multi-ancestry genetic study in 5,876 patients identifies an association between excitotoxic genes and early outcomes after acute ischemic stroke"

### Supplementary Materials and Methods

##### Genotyping Information: Genotyping Batch details

All participants were genotyped using Illumina SNP array technology in seven batches:

1. Illumina Human Omni5 Exome 2v1: 202 individuals were genotyped in 204.
2. Illumina Human Core Exome: 307 individuals from the Finnish cohort were genotyped with this platform in two batches, one in 2015 and another one in 2018.
3. Illumina Human Core Exome 24v1: 889 individuals were genotyped in 2016.
4. Illumina Human Core Exome 12v1: 1,255 individuals were genotyped in 2017.
5. Illumina Human Exome 24v1.2: 1,396 individuals were genotyped in 2018.
6. Illumina Infinium GSA-24 v2.0: 1,923 individuals were genotyped in 2019.

### Supplementary Results

##### Functional Annotation of the remaining Loci

Gene based analyses for 2p25.1 suggested that *RNF144A* was the driving gene in this locus (p=0.035). However, no eQTLs for this gene or others was found in any of the publicly available datasets. MR analyses did not suggest any causal relationship between *RNF144A* and ΔNIHSS. For 2q31.2, no gene reached nominal significance in that locus. However, the top hit rs13403787 was a cis eQTL for *DFNB59* in temporal lobe cortex ($p=6.40\times10^{-05}$) in the Braineac dataset. We evaluated the expression of *DFNB59* in an in-house single nuclei RNASeq dataset derived from the parietal lobes of human brains (http://ngi.pub/snuclRNA-seq/). It was expressed in all brain cells, but especially in microglia ($p=3.54\times10^{-54}$) (Supplementart Figure 4D). The transcript was not present in the GTEx portal dataset and no MR analyses could be conducted to test if there was any causal relationship between *DFNB59I* and ΔNIHSS.
The tagging variant in 4q34.3 locus, rs12641856 was found to be a cis eQTL for *MGC45800* in thalamus ($p=2.20\times10^{-03}$) in the Braineac database. However, this gene was not identified by the gene-based analyses, It was not found to be differentially expressed in brain or in any brain cell subtype, and could not be tested in the GTEX dataset by MR because it was missing in this dataset.
None of our analyses identified any genes or eQTLs for the 6q26 locus. Even though no gene was identified by the gene-based analyses, several eQTLs for *TWISTNB* in cultured fibroblasts (GTEX portal: $p=4.20\times10^{-05}$) as well as *ABCB5* in putamen (Braineac: $p=7.10\times10^{-05}$) and thalamus (Braineac: $p=3.20\times10^{-05}$) were found for 7p21.1. *TWISTNB* was found to be expressed in all cell types from brain cortex, but especially in oligodendrocyte progenitor cells (OPCs) ($p=5.66\times10^{-08}$) (Supplementary Figure 4E). *ABCB5*, does not seem to be expressed in brain cortex according to the single nuclei RNASeq dataset (Supplementary Figure 4F). No MR analyses could be performed because the transcripts were not present in the tested datasets.
